# An Implantable Device that Converses with Patients and Learns to Co-Manage Epilepsy

**DOI:** 10.64898/2026.01.26.26344234

**Authors:** Zack Goldblum, Haoer Shi, Zhongchuan Xu, William KS Ojemann, Carlos A Aguila, Kevin Long, Kevin Xie, Kerry C Nix, Katie Walsh, Ellie Chang, Sarah Lavelle, Brandon Bach, Kathryn A Davis, Nishant Sinha, Lauren H Hammer, Erin C Conrad, Brian Litt

## Abstract

One-third of the world’s 70 million people with epilepsy have seizures that are not controlled by medication; and implantable devices are an exciting option for treatment. These devices improve seizure control and can detect impending attacks, missed medication, and impaired cognition. Unfortunately, they have no way to share this information with their hosts in real-time – a limitation common to most medical devices. This is a missed opportunity for implants and wearables to learn from patients, focus on what matters most to them, and teach them how their behavior affects their health. Here, we present a device platform that converses with patients and learns to co-manage epilepsy. The inpatient prototype links scalp and intracranial EEG (electroencephalograms) to secure large language models that communicate freely and bidirectionally with their hosts through a smartphone app. An AI agent ingests biomarkers of sleep, medication level, cognition, and seizure risk extracted from brain activity. It converses with patients to inform them of clinical events and physiological trends, records their symptoms, responses, and behaviors, and automatically retrains itself to improve performance. Both patients and the AI agent can initiate conversations to teach each other and personalize interactions. We demonstrate this platform in 13 patients undergoing inpatient video-EEG monitoring for epilepsy and validate its performance. Algorithms for detecting seizures optimized their precision over several days without expert intervention – in contrast to the months of iterative, in-person physician programming currently required. Patients responded positively to messages regarding sleep, cognition, and seizure risk while rating the system as highly usable. The platform includes several safeguards, including a system for further algorithm fine-tuning using efficient expert review, and features that ensure data security and regulate communication content. Further work will link other biosensors to measure behavior, improve performance, and optimize therapeutic stimulation. We propose this system as a scalable platform for medical devices that can rapidly adapt to patient and provider needs; one that is broadly adaptable to improving care for many medical conditions.

## Introduction

*A 30 year-old veteran walks into a bar. None of the patrons are aware of the anti-seizure device implanted in his brain, nor the traumatic brain injury that requires it. It’s a hot day, and he quickly downs a cold beer. Within minutes his phone vibrates with a text from his implantable. “What are you doing Dave? Your probability of seizure within the next 6 hours increased to 64%.” “Just had a beer” Dave replies. The device pauses briefly, then resumes, “I’ll remember that. I’d suggest not having another one. I’ll stimulate for now and text if things worsen. Above 80% probability you might consider taking 0.5 mg of Ativan and calling an Uber*.

More than one-third of the world’s 70 million people with epilepsy continue to have seizures despite aggressive treatment with medications. The cost of this individual and societal burden is enormous, as measured by economic impact, longevity, and impaired quality of life [1]. Some medication-resistant patients will be candidates for functional surgery – removing or laser-ablating seizure-generating brain regions – but only half will become seizure-free two years after surgery [2]. Implantable devices are an exciting, less invasive alternative to these procedures. They regularly improve seizure control without surgery’s most severe side effects, such as cognitive impairment or mood disorders. However, seizure-freedom is rare with devices.

Even for the two-thirds of patients with medication-responsive epilepsy, living “seizure-free” but reliant on medication presents its own challenges. These individuals are at risk for seizure relapses triggered by common life events: forgetting or running out of medications, sleep deprivation, pregnancy, or taking commonly prescribed medications that increase seizure risk. There is a need for technologies that help people *live* with epilepsy, such as the fictional partnership between Dave and his device, because clinical treatment often fails to provide complete seizure freedom.

## Epilepsy devices

There are currently only three devices approved by the FDA to treat epilepsy. These devices are either open-loop, meaning they stimulate their targets in on/off cycles agnostic to neural activity, or closed-loop – delivering stimulation in response to pre-programmed detection criteria while monitoring brain or other activity. These devices reduce seizures gradually over years, plateauing at a median seizure reduction of 75% nine years after deployment in the most positive studies [3], though approximately 30% of patients have minimal or no improvement. While some devices can store and transmit small amounts of data asynchronously, none can share it in real-time. This is a missed opportunity to share useful information with patients which the device “knows” but has no way to convey. This information could include changes in seizure risk, the presence of known seizure precipitants, or recognizing specific patient behaviors that may be either beneficial (e.g., getting more sleep) or detrimental (e.g., consuming alcohol or some medications) to controlling seizures. Similarly, current devices deprive patients of the opportunity to configure them to their needs and lifestyle, annotate collected data, and efficiently re-train algorithms with minimal physician interaction.

New minimally-invasive EEG devices are poised to dramatically change seizure management with their ability to continuously collect data from patients during normal daily life. These devices vary from small, battery-powered single-channel recording motes affixed to the scalp with an adhesive, to subgaleal wires and miniaturized electronics placed under the scalp, to wearable devices [4]. Some devices can transmit continuous EEG to smartphones and subsequently to the cloud, where the data is processed and viewable by clinicians. Wearable devices monitor movement, heart rate, and other peripheral parameters to alert families to convulsive seizures. Unfortunately, none of these devices have bi-directional interfaces for patients to communicate and interact with them in real-time.

Based upon our experience and discussions with clinicians, caregivers, and patients, we propose that an optimal interactive epilepsy device should have the following features: (1) the ability to detect, process, and present information transduced from sensors, in this case scalp or intracranial EEG (iEEG), and present it to patients in an intelligible fashion, (2) the ability to respond to patient queries with actionable answers and convey information in text, figures, and modalities that adjust to the patient’s level of function, (3) an intuitive, conversational chat interface with minimal latency that can deliver patient surveys and facilitate two-way, natural conversation, either through voice or text, (4) the ability to use patient responses to annotate data and re-train algorithms that dynamically adapt to patient needs (e.g., training a seizure detector), (5) governance functions that monitor the appropriateness and truthfulness of output to patients and guarantee safety and information security, and finally, (6) it was strongly felt that experts – physicians and other providers – must have the ability to efficiently view and annotate data, both for patient care and to improve device performance. Expert annotations, in addition to patient annotations, are necessary to compensate for when patients are amnestic to seizures or in cases where their annotations may be less reliable. The platform also needs to adapt to different patient needs, conditions, and care environments, such as the hospital, outpatient clinic, and home. Computation, data transmission, and storage must be cost-efficient and scalable. Optimizing resource utilization and scalability were not the focus of this proof-of-principle study, but are considered in the Discussion below.

In the following sections, we describe a platform that performs these functions, and report our results from testing it prospectively on 13 patients admitted to the Hospital of the University of Pennsylvania epilepsy monitoring unit (EMU) for continuous video and scalp or iEEG monitoring.

## Results

### Patient recruitment

We conducted a prospective, single-center observational study to evaluate a real-time patient–data interface, including its clinical utility and usability; detailed study procedures are described in the Methods. Thirteen adult EMU inpatients undergoing continuous EEG monitoring for seizure characterization as part of clinical management or presurgical evaluation were recruited (scalp EEG n = 11; iEEG n = 2; see Table 1 for cohort demographics and Fig. S1 for the patient flow diagram). Patients were enrolled during their clinical admission without altering standard-of-care monitoring or treatment. Throughout their EMU stay, participants were encouraged to interact with the platform and provide feedback on their experience.

**Table 1:**
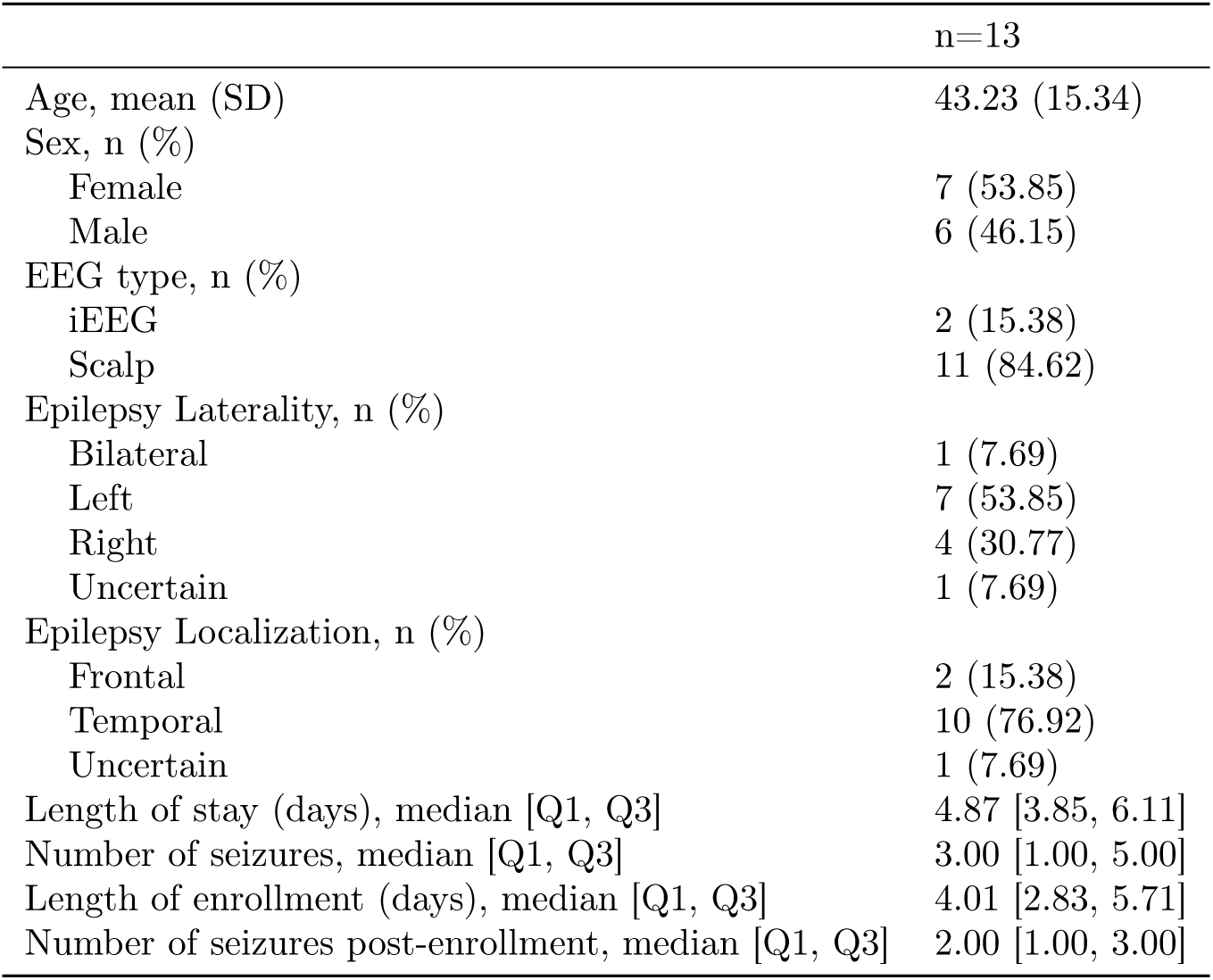
Patient demographics.

### Device platform

The study employed a novel platform that comprises three systems: (1) software that extracted real-time EEG and metadata from the hospital network, (2) a HIPAA-compliant cloud analytics environment that analyzed this data stream, and (3) an app installed on the patient’s smartphone that used an integrated conversational agent to facilitate bidirectional communication. The agent spontaneously messaged patients to deliver clinical information, alerts, and surveys, while patients used the app to query their physiological data, understand trends, and provide behavioral annotations regarding their symptoms to inform seizure-detection algorithms. The platform is described in detail in Fig. 1, below.

**Figure 1:**
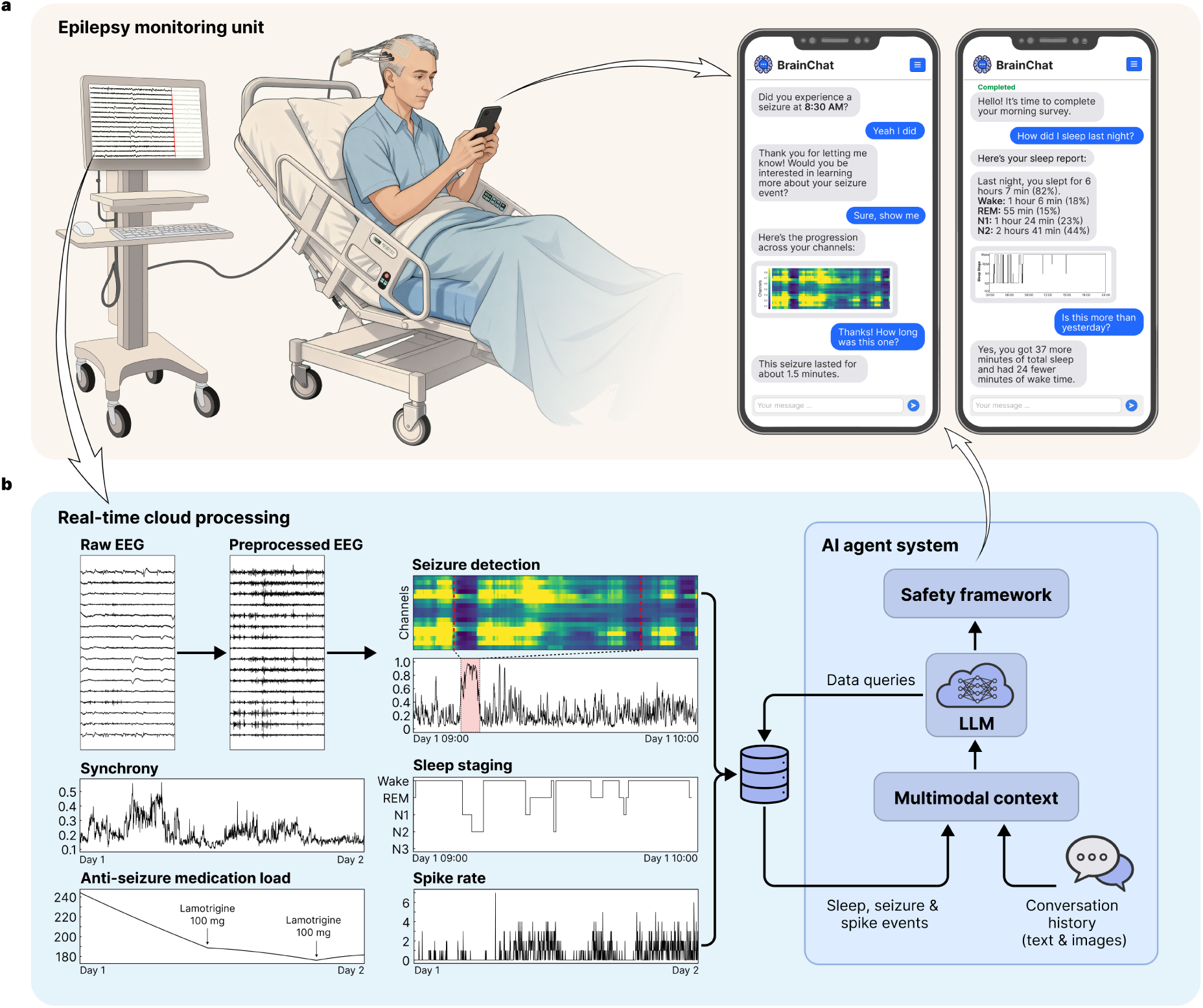
Platform overview. **a.** Patient-AI interface in the epilepsy monitoring unit (EMU). *Left:* Patients undergoing con-tinuous scalp or intracranial video-electroencephalography (EEG) monitoring connect to the patient-data interaction platform via a smartphone application. EEG data streams from the EMU to a secure cloud environment for real-time processing. *Right:* Representative screens show bidirectional interaction modes, including system-initiated alerts (seizure alarms, surveys) and patient annotations. The interface delivers neurophysiological visualizations, such as seizure heatmaps and sleep hypnograms, directly within the chat stream. **b.** Cloud processing and AI agent system. *Left:* Real-time neurophysiological biomarkers extracted from EEG, including seizure probability, sleep staging, spike rates, and brain synchrony, are compiled in a structured database. *Right:* The large language model (LLM)-based agent system manages patient interactions and queries this database to synthesize responses.

The patient was sent surveys three times per day – morning, afternoon, and evening – consisting of 10–15 questions asking about mood, anxiety, sleep, energy level, and overall well-being (complete survey content is detailed in the Supplemental Materials). The real-time EEG recorded from patients in the EMU was streamed from the hospital network into the cloud infrastructure where feature extraction and inference were performed using pretrained models, including SPaRCNet [5], WaveNet [6], and ONCET (unpublished work) for seizure probabilities; SpikeNet [7] and customized intracranial EEG spike detection algorithm [8] for the times, locations, and rates of interictal epileptiform discharges (spikes); and YASA (Yet Another Spindle Algorithm) [9] for sleep staging. Additional features included phase synchrony (a biomarker for anti-seizure medication load [10]), and anti-seizure medication load estimated using the medical records. The rationale behind these measures is that they demonstrate the platform’s ability to extract information that can correspond to seizures, clinical events, sleep architecture, and warn of changes in medication level that can identify increased risk of symptomatic seizures or convulsions. For instance, post hoc analyses using a linear mixed-effects model revealed a significant inverse relationship between phase synchrony and medication load (correlation coefficient −0.315, 95%CI −0.337 to −0.293), indicating that synchrony increases as medication load decreases (Fig. S2). To enable patient interaction with these data, the platform utilized two distinct AI agent modes: ‘general chat’ for conversational flow and subjective logging, and a ‘data agent’ that could synthesize and retrieve the aforementioned neurophysiological features from the study database. Patients were able to ask questions about these data, and request answers as text or figures. HIPAA-secure instances of GPT-4o-mini (OpenAI, San Francisco, CA) were used in the AI agent system for communication between the patient and their device. Fig. 2, below, depicts the platform architecture, including the AI agent system implementation. These are explained in greater detail in the Methods section.

**Figure 2:**
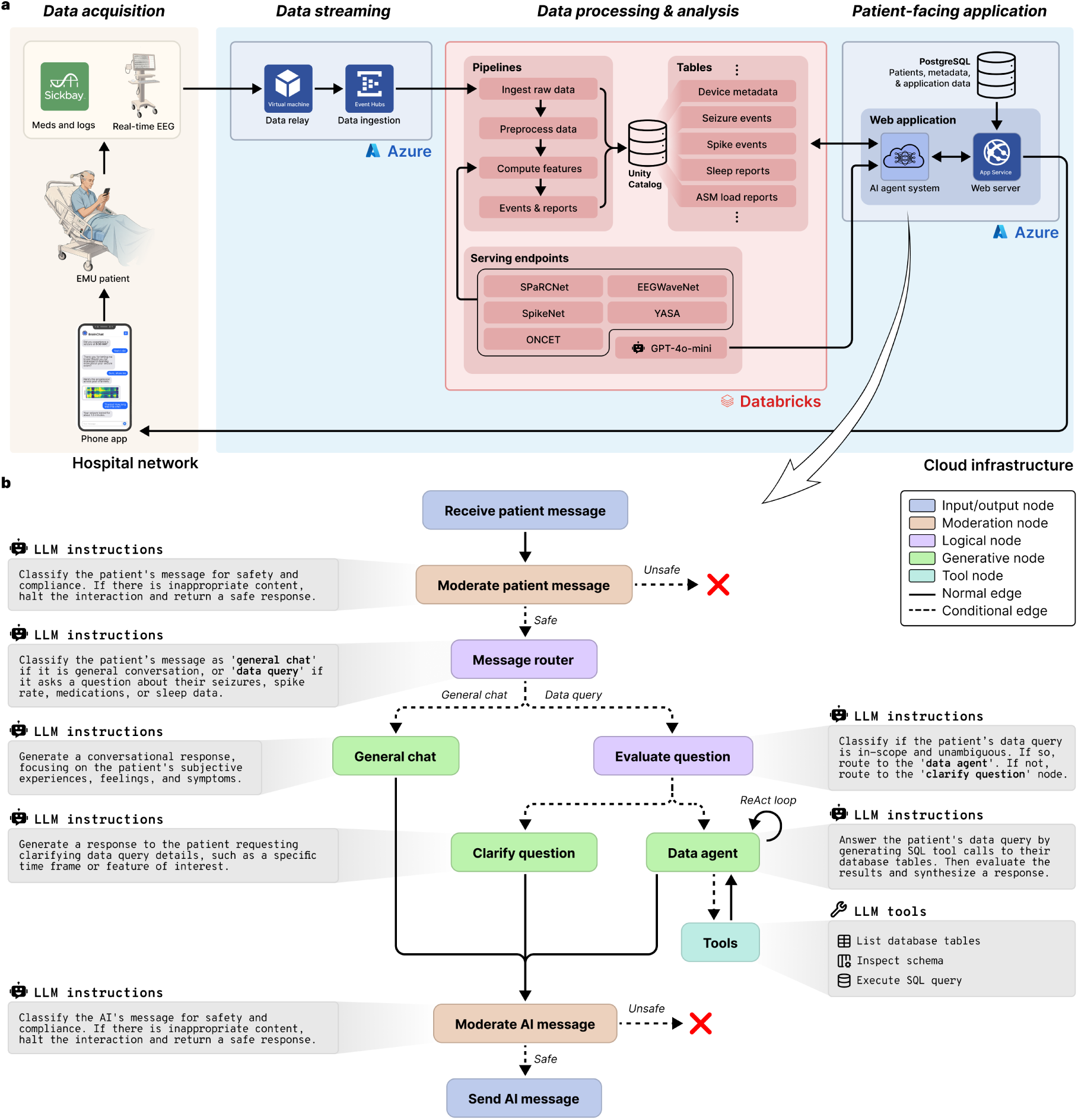
Platform technical architecture and AI agent framework. **a.** Cloud infrastructure and real-time data streaming. The platform integrates on-premises clinical data acquisition with HIPAA-compliant cloud services. Continuous EEG signals are acquired via clinical workstations and relayed through a cloud-hosted virtual machine to Azure Event Hubs for ingestion. Simultaneously, clinical metadata, including medication dosing and logs, are retrieved via the Sickbay platform. Data is processed within a Databricks environment using Delta Live Table pipelines for preprocessing, feature extraction, and real-time clinical event detection. Automated analyses are performed by dedicated serving endpoints that host deep learning models and other algorithms. A HIPAA-secure version of GPT-4o-mini is utilized in the AI agent system. **b.** Directed state machine architecture of the AI agent system. All incoming patient messages and outgoing AI responses pass through moderation nodes that evaluate safety and clinical compliance. A message router classifies inputs as either ‘general chat’ for regular conversation or ‘data query’ for clinical information retrieval. Data queries are subsequently evaluated for scope and ambiguity. Valid queries are handled by a data agent employing a ReAct (reasoning and action) loop. This agent autonomously invokes SQL tools to inspect database schemas and execute secure queries against the Unity Catalog, where patient data is stored. The agent maintains a persistent state, including conversation history and multimodal context (text and images) for context-aware interactions.

### Patient engagement

We analyzed the 1,307 messages exchanged between the 13 patients and the system over the study period (Fig. 3a) to evaluate its ability to engage patients and facilitate real-time co-management of epilepsy. The median patient participation, due to limited time in the EMU, was 4.85 days (IQR 3.85–6.11; Table 1). The system initiated 68.9% (n=900) of interactions, primarily consisting of general chat messages (43.1%), scheduled surveys (13.7%), and clinical event notifications (10.3%). Patients initiated the remaining 31.1% (n=407) of interactions. Patient engagement was predominantly conversational, with 88.5% (n=360) of patient-initiated messages categorized as general chat, indicating that patients were comfortable using the platform for regular conversation and subjective logging. Patients also utilized it for clinical data retrieval, directing 11.5% of their queries toward insights about seizure events (5.9%), sleep metrics (3.7%), and interictal spike rates (1.9%).

**Figure 3:**
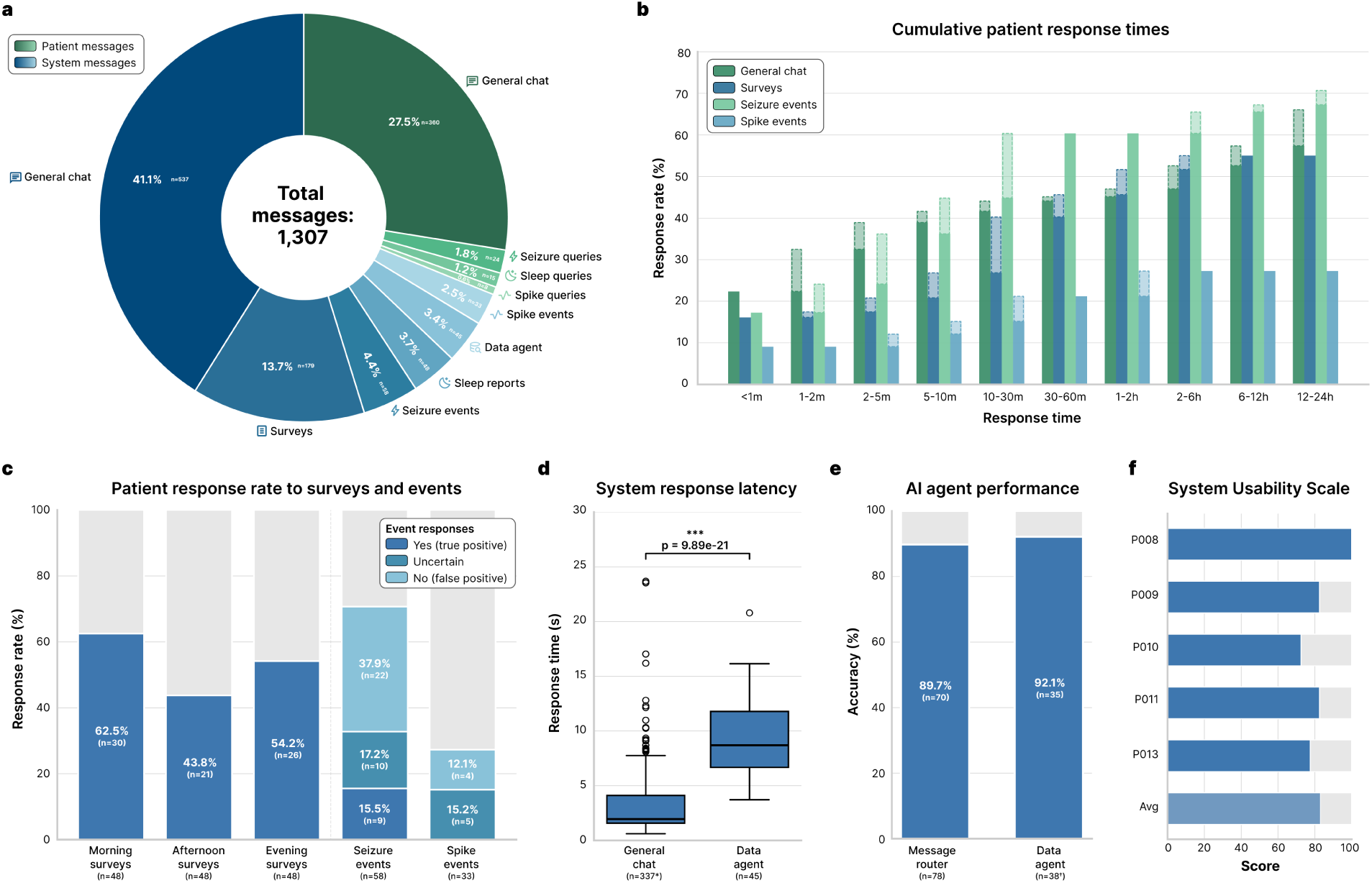
Patient engagement and technical performance of the AI agent system. **a.** Distribution of total messages (n=1,307) exchanged between patients (n=13) and the system, stratified by initiator (system vs. patient) and interaction category. **b.** Grouped bar plots illustrating the percentage of patient responses received across progressive time intervals for four categories: general chat, surveys, seizure events, and spike events. The total height of each bar represents the cumulative response rate up to that timeframe; the lighter, dashed regions denote the marginal increase in the response rate added at each time interval. **c.** Survey response rates and EEG event validation. *Left:* Bar plots showing patient response rates for morning, afternoon, and evening surveys (n=48 each). *Right:* Stacked bar plots for real-time seizure (n=58) and interictal spike alerts (n=33), showing both the response rate and patient-provided annotations (‘yes’/true positive, ‘uncertain’, or ‘no’/false positive). **d.** Response latency comparison between the general chat and data agent nodes. Box plots show median and interquartile range (IQR); whiskers extend to 1.5×IQR. The data agent exhibited significantly higher latency due to its reasoning-action framework and increased computational overhead (p=9.89×10^-21^, two-sided Mann-Whitney U test). **e.** AI agent performance. *Left:* Message router accuracy in directing queries (n=78) to either the general chat or the data agent node based on patient intent. *Right:* Data agent accuracy in executing retrieval and analysis tasks (n=38) on patient data. **f.** System Usability Scale scores reported by patients (n=5). The distribution places the system in the "excellent" usability range (mean score 83/100). *200 automated (non-AI-generated) general chat messages were excluded from the latency analysis. ^†^7 patient general chat messages erroneously routed to the data agent were excluded from the data agent accuracy analysis.

To characterize the temporal dynamics of patient engagement, we computed the cumulative response times across interaction types (Fig. 3b). Patients exhibited distinct response patterns depending on the urgency of the interaction. Seizure queries elicited the most rapid engagement, with 44.8% of responses occurring within 10 minutes and response rates reaching 60.3% within the first 30 minutes. Conversely, notifications for spike events elicited the lowest engagement, plateauing at 27.3% within the first hour. This likely reflects the lower patient prioritization of biomarkers that are often asymptomatic and imperceptible. Survey response rates reached 55.0% before their 3-hour expiration time. General chat interactions continued to accumulate steadily over the full 12–24 hour window, suggesting that patients treated these interactions as an asynchronous task compatible with the routine activities of the inpatient environment.

Patient-AI interactions enabled patient-in-the-loop validation of automated EEG detections. Patients demonstrated high engagement with high-priority alerts; the response rate to real-time seizure detections (n=58) was 70.6% (Fig. 3c). Patients confirmed 15.5% (n=9) of these alerts as true seizures, rejected 37.9% (n=22) as false positives, and labeled 17.2% (n=10) as uncertain. All uncertain events (n=9) were marked as non-seizure by experts. This provided a source of discriminatory labels for algorithm fine-tuning that is absent in passive monitoring. Patients were also asked about symptoms related to spike events, such as cognitive difficulties, which have been described in prior studies [11]. Patient engagement with sub-clinical interictal spike notifications (n=33) was markedly lower, with a 27.3% response rate and zero patient-confirmed events. This is not surprising, as patients are rarely aware of interictal discharges, though they can have negative effects on cognition. Adherence to surveys varied by time of day, with the highest response rates observed for morning surveys (62.5%, n=30), likely driven by the subsequent delivery of the daily sleep report, compared to afternoon (43.8%, n=21) and evening (54.2%, n=26) surveys.

The system’s technical performance was evaluated for its two primary modes of interaction: ‘general chat’ and ‘data agent’ (Fig. 3d). We observed a statistically significant difference in response latency between these modes (Mann-Whitney U=1086.0, p=9.89×10^-21^; Fig. 3d). General chat operated with near real-time fluidity (median latency: 1.95 s; IQR 1.57–4.11 s), whereas the data agent, which required dynamic SQL generation and database querying, exhibited higher latency (median: 8.69 s; IQR 6.67–11.79 s). Despite the increased complexity of data retrieval tasks, the AI agent system maintained high reliability. The message router correctly classified and directed patient intents to the general chat or data agent with 89.7% accuracy (n=70/78), and the data agent successfully executed retrieval tasks with 92.1% accuracy (n=35/38; Fig. 3e). The AI agent system effectively moderated patient-AI interactions within the clinical environment. The system flagged and blocked 1.47% (6/407) of patient messages for inappropriate or unsafe content and prevented them from reaching the LLM. Although, upon post hoc human review, the messages were only flagged due to lack of context handling in an early iteration of the moderation system, or due to an auto-correct error in one case. None of the patient messages were intentionally harmful. The AI output moderation layer flagged 0.0% (0/908) of LLM-generated messages, indicating that the system did not generate any unsafe or inappropriate messages during the study period. These results were also corroborated by human review.

Patient feedback regarding the system’s usability was very positive. The System Usability Scale (SUS) [12] scores indicated a high degree of satisfaction, with a mean score of 83/100 (n=5; Fig. 3f). Individual scores ranged from 72.5 to a maximum of 100, placing the system’s usability in the "excellent" range of the SUS spectrum and demonstrating that the chat-based interface was accessible and intuitive for patients in an acute hospital setting.

### Seizure detection

Accurate seizure detection is an essential feature for seizure monitoring devices. Nevertheless, current algorithms, while demonstrating promise in specific applications, do not generalize well [13]. We implemented rolling fine-tuning using real-time patient feedback on seizure detections as weak labels to improve seizure detection without requiring clinician input (Fig. 4). To set a baseline, we evaluated previously validated models – SPaRCNet for scalp EEG and ONCET for iEEG – against technician- and clinician-annotated seizure events. The non-fine-tuned models had a median event-level sensitivity of 0.83 (IQR 0.35–1.00; n=11) and a false alarm rate of 0.58 per hour (IQR 0.35–0.77; n=13; Fig. S3). We first performed an offline validation using patient-initiated push-button seizure alarms as annotations, which are standard in the EMU and available for all patients. We treated the window from one minute before to two minutes after each alarm as seizure, and all other intervals as non-seizure (Fig. 5b). These push button events showed a median sensitivity of 1.00 (IQR 0.51–1.00; n=11), a median precision of 0.50 (IQR 0.30–0.66; n=11), and a median F1 score of 0.55 (IQR 0.43–0.72; n=10). Models were updated sequentially every three hours if pre-defined criteria were met (see Methods; Fig. 5a). Each iteration was evaluated on the remaining unseen data, with performance shifts measured against the baseline. We then calculated the median intra-patient improvement from baseline across all fine-tuned iterations. Across patients, fine-tuned models achieved a median 77.8% reduction in false alarms per hour (IQR 40.0–83.3%; median difference −0.305; Wilcoxon signed-rank test, W=3.0, p=0.001; Fig. 5b) and a 115.4% increase in F1 score (IQR 0.0–200.0%; median difference 0.017; Wilcoxon signed-rank test, W=28.0, p=0.008; Fig. 5b). Simulating prospective deployment of the fine-tuned models, the false alarm rate across the EMU stay decreased by 77.3% (IQR 52.0–82.7%; median difference −0.292; Wilcoxon signed-rank test, W=87.0, p=0.001) and the F1 score increased by 66.2% (IQR 0.0–199.1%; median difference 0.109; Wilcoxon signed-rank test, W=0.0, p=0.002), while maintaining baseline sensitivity. Next, we fine-tuned our models using prospective annotations collected through our system. The seizure annotation feature was implemented for the nine most recent participants. Six of these patients had confirmed events (true seizures or false positives), with a median sensitivity of 0.00 (IQR 0.00–0.00; n=6) and a precision of 1.00 (IQR 0.50–1.00; n=6). After fine-tuning with this data, we achieved a 73.4% reduction in false alarms (IQR 16.4–92.1%; median difference −0.451; Wilcoxon signed-rank test, W=4.0, p=0.109; Fig. 5c) and a 159.1% increase in F1 score (IQR 29.5–352.5%; median difference 0.111; Wilcoxon signed-rank test, W=15.0, p=0.031; Fig. 5c). Across the EMU stay, the decrease in false alarm rate and increase in F1 score were 76.3% (IQR 16.9–81.0%; median difference −0.459; Wilcoxon signed-rank test, W=18.0, p=0.078) and 190.3% (IQR 22.2–308.3%; median difference 0.131; Wilcoxon signed-rank test, W=1.0, p=0.031), respectively.

**Figure 4:**
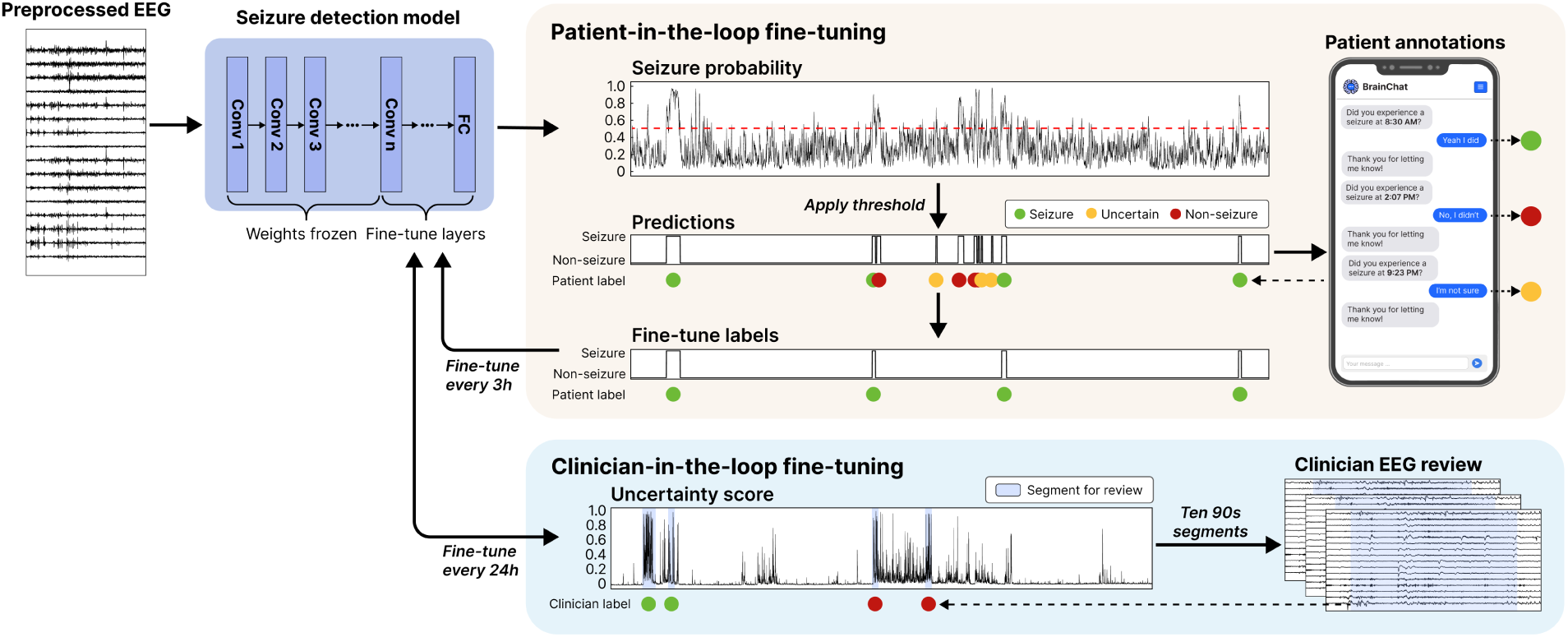
Workflow for fine-tuning of seizure detection models. Pretrained seizure detection models produced seizure probability time series from preprocessed EEG windows; thresholds were then applied to generate binary predictions and identify discrete seizure events. Upon detection of a seizure event, a push notification was sent to the patient for confirmation, allowing responses of ‘yes’, ‘no’, or ‘uncertain’. These responses were converted into labels for fine-tuning: seizure annotations were labeled as 1, uncertain annotations as NaN, and all other labels as 0. Models were fine-tuned every 3 hours, with only the last few layers trainable while all other layers remained frozen to prevent catastrophic forgetting – abruptly forgetting previously learned information after being trained on new data. To further improve performance, a clinician-in-the-loop approach was implemented every 24 hours. Ten 90-second EEG segments with the least confident predictions were selected and reviewed by clinicians for fine-tuning.

**Figure 5:**
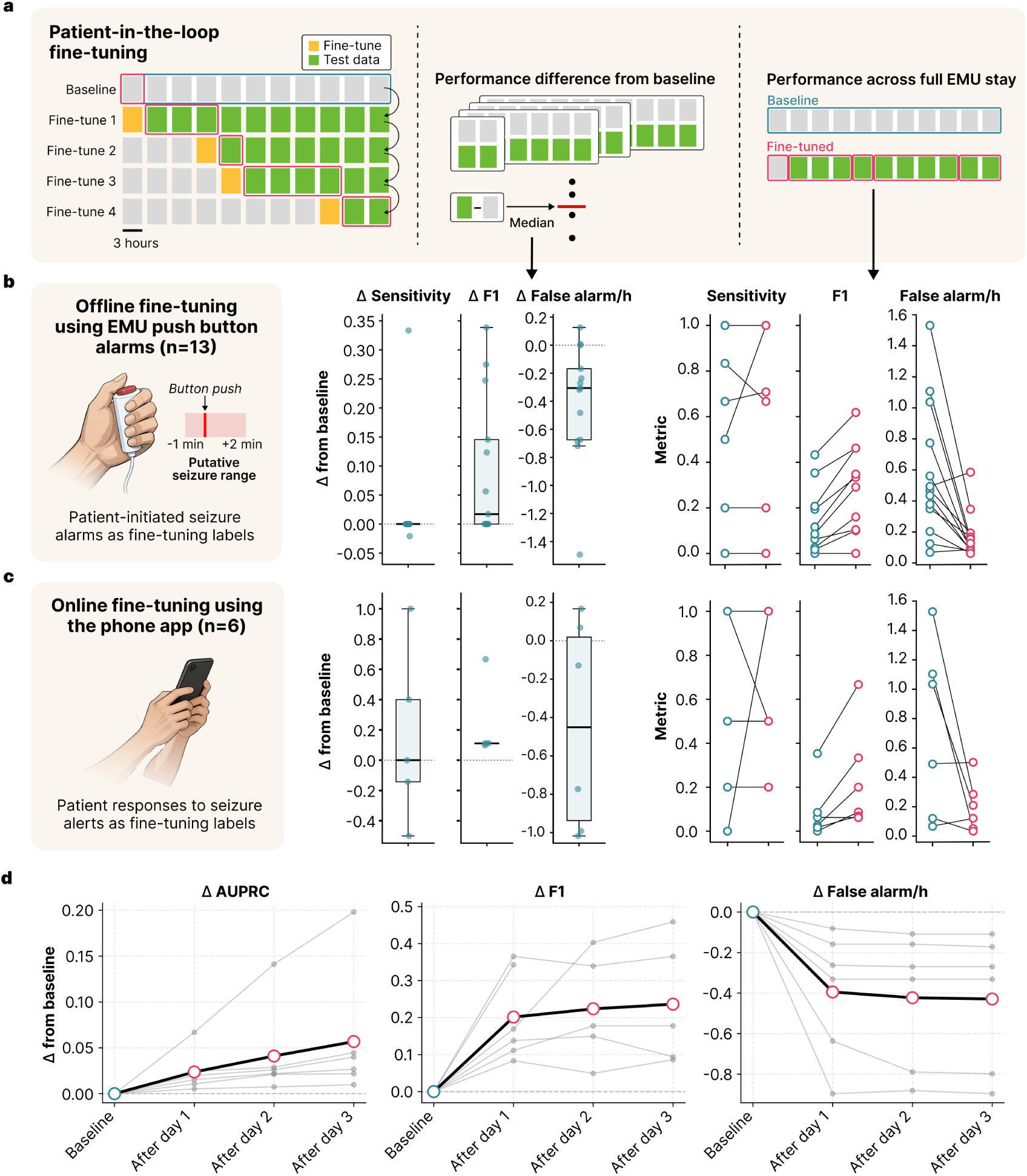
Patient- and clinician-in-the-loop fine-tuning improves seizure detection. **a.** Evaluation strategy for patient-in-the-loop fine-tuning. *Left:* The baseline model was fine-tuned sequentially every three hours when predefined criteria were met. Each fine-tuned model was evaluated on the remaining test data and compared with the baseline model. *Middle:* For each patient, the median performance difference from baseline was calculated across fine-tuning iterations. *Right:* Performance across the full EMU stay under prospective deployment of the fine-tuned models was also compared with baseline. **b.** Fine-tuning performance in offline validation using patient-initiated push-button alarms as labels. **c.** Fine-tuning performance using patient responses to seizure alerts through the phone app as labels. **b.** and **c.** *Middle*: Median change from baseline in seizure event sensitivity, F1 score, and false alarms per hour after fine-tuning. Each dot represents one patient. Boxes indicate the interquartile range (IQR) with the central line representing the median across patients; whiskers extend to 1.5×IQR. *Right* : Paired baseline versus fine-tuned performance across the EMU stay. Blue and red dots indicate baseline and fine-tuned performance, respectively, for each patient. **d.** Change in seizure detection performance after clinician-in-the-loop fine-tuning. Gray lines indicate individual patients and the black line shows the median change across patients. AUPRC: area under the precision–recall curve.

We also implemented a clinician-in-the-loop framework in which the system automatically flags uncertain predictions and requests clinician annotations, providing an additional layer of verification to prevent degradation of model performance during real-time fine-tuning. In offline validation, clinician-in-the-loop fine-tuning with EEG technician annotations, another source of validated markings, increased F1 score by 271.5% (IQR 111.4–344.7%; median difference 0.185; Wilcoxon signed-rank test, W=21.0, p=0.016), increased in AUPRC by 89.3% (IQR 11.6–138.7%; median difference 0.042; Wilcoxon signed-rank test, W=21.0, p=0.016) and decreased false alarm rate by 86.6% (IQR 67.9–90.9%; median difference −0.284; Wilcoxon signed-rank test, W=0.0, p=0.031) (Fig. 5d).

## Discussion

In this study, we demonstrate a novel approach to medical devices: a platform that interprets physiological activity and behavior in real-time and allows two-way, spontaneous communication between patients and their devices to co-manage disease. Utilizing inpatient EEG monitoring equipment, together with compute, storage, and LLMs in HIPAA-secure cloud environments, patients communicated with their devices through a smartphone application. They were able to initiate conversations, receive alerts, ask questions to query their data, and adjust its behavior, including conversation content. Despite the limited setting and short duration of participation, patients found this platform to be useful in better understanding and managing their condition, with five of the 13 spontaneously requesting access to it as outpatients. The system learned and re-trained itself to markedly improve seizure detection performance using imperfect patient annotations alone, without the need for clinician intervention – though time-efficient expert annotation further improved performance. System latencies were deemed acceptable by this small patient cohort: response rates were approximately two seconds for simple conversational interactions and nine seconds for complex queries that synthesized graphs and data in addition to text.

The clinical and scientific impact of this work is multifold.

1. **A new approach to disease management.** Some of the most debilitating aspects of episodic conditions like epilepsy are the unpredictability of events and managing risk, even when patients are doing well. Current EEG devices can detect biomarkers of increased seizure risk, which are often precipitated by patient behavior, such as missed seizure medication, severe stress, sleep deprivation, a new prescription (e.g., antibiotics or antidepressants), or drug or alcohol ingestion. If patients are alerted to these neurophysiologic changes in time, it is often possible to take action to reduce their acute seizure risk by taking additional or a rescue medication [14]. A pilot study, performed in Australia, implemented a seizure warning system that reduced seizures, lowered medication burden, and improved quality of life in a group of 15 patients where iEEG was monitored with a brain implantable coupled to belt-worn warning device [15, 16]. In this study, patients received feedback that was not immediate, but “smoothed” into 30–60 minute time windows, in the form of an illuminating light: red (warning), blue (safe), or white (uncertain). There was no capacity to give patients immediate feedback or tie changes in physiology to behavior. Our study builds upon this foundation, with the capacity for more explicit and immediate feedback in both directions. In the Australian study, algorithms had to be manually retrained at intervals typically months apart. In contrast, our system automates model fine-tuning every three hours, with patients providing annotations directly. Although annotation inaccuracies may arise from missed or delayed responses, postictal amnesia preventing patient input, or subclinical seizures that go unrecognized, these imperfect labels nonetheless proved effective for fine-tuning, consistent with prior studies demonstrating the utility of weak labels in model training [17, 18]. We additionally implemented a clinician-in-the-loop strategy that allows experts to efficiently review large amounts of preprocessed data to validate and improve upon weak annotations from patients. Our group has previously validated this approach using data from implanted, closed-loop devices, demonstrating that expert review of only a small fraction of EEG epochs is sufficient for accurate clinical classification of thousands of events [19]. The clinician-in-the-loop system serves multiple purposes: (1) to validate the reliability of patient markings and detections, particularly for events when the patient was not able to answer, (2) to refine system performance above that possible from patient markings alone, when possible, and (3) to assess the need for expert algorithm supervision for specific patients. This last function will be important to scaling the platform for use in the outpatient setting, as once the system is trained and validated it may need only rare expert review, greatly reducing resource requirements. While it is true that many well-controlled patients may not want to be bothered by the presence of a continuous monitoring device reminding them of their epilepsy, our discussions with patients indicate that most individuals with epilepsy experience time periods when this information could be invaluable, such as when trying to become pregnant (medication levels may drop precipitously), during a systemic illness, or when changing, increasing, or tapering medications. The ability to rapidly adapt systems like ours could make these systems easy to use and available on demand. For example, this might be done by setting alert thresholds for events patients deem vital to know, turning them on or off as needed just by speaking to them, or by removing a wearable sensor or external transmission device when its function is not desired.

2. **An expanding platform.** The innovations we present are not limited to epilepsy, and are easily implemented broadly where patient communication can enhance the efficacy of medical devices. For example, we are currently readying a trial of our platform for programming deep brain stimulation (DBS) devices in patients with Parkinson’s disease. Rather than iteratively visiting a doctor’s office every month after implant for expert programming, we propose that an efficient algorithm for searching the stimulation parameter space coupled to two-way patient communication could be a more efficient and less costly method for optimizing tremor and stiffness in an ecologically valid environment. We imagine a simple interaction, where the device asks the patient if they feel their gait is adequately treated, and then the patient goes for a walk while parameters are systematically adjusted. Coupling these algorithms to external sensors that transduce movement will add to device precision, particularly if motor symptoms limit interaction with a hand-held device. Such a system could substantially expand access to DBS therapy, which is currently limited in part by access to movement disorders specialists that program these devices [20]. Other applications are feasible, such as cardiac devices that alert patients to behaviors which adversely affect heart rhythm or ischemia, and insulin pumps that educate patients about how particular foods or medications affect their serum glucose, and allow them to query their device about metabolic status when they don’t feel well. An exciting frontier in this space is devices to treat neuropsychiatric disorders, such as depression, obsessive-compulsive disorder, cognitive dysfunction, Tourette’s syndrome, addiction, and others. With these conditions, external sensors may not be well-suited to detecting a patient’s inner symptoms or signs, but nuances in communication with an AI agent, in addition to patients expressing their feelings and symptoms, could radically improve the effectiveness and personalization of these devices.

3. **Variable connectivity.** This study could extend to the outpatient setting with a design that distributes processing tasks between a smartphone and the cloud. In this model, basic functions like seizure detection and warnings would be run locally by low-overhead agents that leverage dedicated neural processing units in modern smartphones. This allows for the use of quantized language and deep learning models to ensure continuity of care regardless of internet connectivity [21, 22]. Computationally intensive tasks, such as model re-tuning, could then be prioritized for the cloud when connection is available. This hybrid architecture is particularly practical for low-channel-count hardware, such as recently FDA-approved subgaleal EEG systems, where reduced data dimensionality simplifies on-device inference. Crucially, as we have shown that algorithm improvement is possible using patient-derived annotations, we anticipate that these systems will be able to individualize and improve performance even when disconnected from the web or expert intervention.

### Limitations of our results

Our results report on a small group of 13 patients admitted to the EMU for evaluation. We chose this group because of the proximity to secure computing resources, high-quality neurophysiology data, and continuous supervision of patients with video and multiple sensors to ensure that LLM-based agents could do no harm to patients. We used both scalp and intracranial EEG patients to demonstrate generalized use of our platform, and that it could operate equally well on both signal modalities. While we recorded continuous video, electrocardiogram, and data from other sensors used in routine epilepsy monitoring, these data were not utilized in these first proof-of-principle experiments. Patient surveys and communications were carefully monitored for inappropriate content, and after analysis, none was found (see Results above). Several patients declined to participate in our study due to a lack of desire to participate in research, or because they had agreed to other research investigations they wished to prioritize while admitted to the EMU. This may have introduced some selection bias for individuals more interested in AI and technology, improving our patient satisfaction scores. Response rates, while considered good for survey studies, were still below 100%, which could also have affected our results. It is anticipated that when this platform is deployed in the outpatient setting, the frequency of patient-device communication will fluctuate over time, as patients personalize their devices according to features they find most valuable. Unlike existing warning systems that rely on expert-adjusted thresholds, we anticipate that patients will be able to individualize their thresholds for warnings and interactions to maximize benefit and minimize alarm fatigue. The seizure annotation feature was not implemented for the first four participants and was subsequently developed and iteratively refined based on patient feedback. We also introduced several improvements to mitigate initially low response rates, including adjustments to the thresholds and criteria for triggering notifications, and displaying alert messages in a pop-up window rather than in-line with other messages. These modifications improved the response rate and annotation accuracy in later participants.

Fine-tuning substantially reduced false alarm rates and resulted in significant improvement in overall seizure detection performance. Detection sensitivity remained comparable to the baseline model and did not improve with patient-specific fine-tuning, likely due to several factors. First, the relatively modest size and homogeneity of the fine-tuning dataset may have been insufficient to reshape the learned feature space of a model pretrained on hundreds or thousands of patients. Second, we adopted conservative fine-tuning hyperparameters, including low learning rates, limited training steps, model distillation, and partial layer freezing to mitigate overfitting and catastrophic forgetting. However, these constraints also limited the magnitude of gradient updates, preventing substantial departure from the pretrained prior. Collectively, the pretrained model had already converged on a robust internal representation of ictal dynamics, with fine-tuning acting as a recalibration mechanism to reduce false alarms. Further adjustment of fine-tuning hyperparameters and investigation into fine-tuning strategies, including synthetic data augmentation and low-rank adaptation (LoRA), may be necessary to achieve additional performance gains, particularly in sensitivity. The seizure detectors employed in this study were selected because of their validated detection performance, lightweight architecture amenable to real-time deployment, and reliable probability calibration. However, they do not reflect state-of-the-art performance for detecting seizures [23, 24]. Adopting a better-performing baseline model would almost certainly improve overall performance. For example, a highly sensitive model that minimizes missed seizures could be fine-tuned to reduce false alarms. We are currently investigating deploying more robust seizure detection models for our application, recognizing that there is rapid innovation in this area, including recently published foundation models [25].

### Next steps and future vision

The true impact of the paradigm we present lies in applying this technology in the outpatient setting to improve everyday life. Proof of real value will come from specific applications and demonstrating clear benefit to patients, caregivers, providers, and insurers in the form of total value care. While teaching patients to identify behaviors or interventions that increase the risk of seizures or adverse clinical events will improve their health, the burden of proof for these outcomes is significant. Similarly, empowering patients to better control their devices and training algorithms with a reduced need for expert interaction will require concrete evidence of benefit to drive widespread adoption. Ideally, future studies will demonstrate reduced seizure frequency or severity, fewer emergency room visits, decreased resource utilization, and improved patient satisfaction. The same potential holds for other domains, such as brain network disorders, heart disease, and diabetes. It may be that informatics approaches – comparing patients equipped with these "intelligent" devices to historical control data gleaned from electronic health records using natural language processing – will hasten this path. If not, controlled clinical trials will be required. The regulatory pathway to approving such enhanced devices will likely be stepwise and gradual, eventually rising to reflect their overall value. Much as autonomous vehicles have undergone a long history of incremental innovation, so may semi-autonomous medical devices evolve, beginning with low-level advisory systems before flourishing into sophisticated clinical partners for diagnosing, managing, and predicting disease.

### Conclusion

In this study, we present a vision for medical devices that freely converse and interact with their hosts to manage disease and improve quality of life. The components to accomplish this are already available, though the algorithms need to be refined and the technology must be made affordable and scalable. With the current pace of innovation in computing, deep learning, and neuroscience, it is only a matter of time before these kinds of interactions become commonplace. The science fiction literature is full of examples of the tremendous potential of joining humans and intelligent devices, but it is also replete with cautionary tales. In the course of our experiments, it was not lost upon us that our early implementation of AI agents in this experiment closely resembled Isaac Asimov’s 3 laws of robotics: (1) that the device can never harm a human through action or inaction, (2) that it must always obey its host except for when it conflicts with the first law, and (3) the device must protect its own existence, as long as this does not conflict with laws 1 and 2. We mention this to acknowledge that others have given careful thought to the challenges and potential consequences of uniting humans with machine intelligence in an intimate way. We believe that because of the great promise of this path, it is important that we push forward rapidly, but acknowledge that progress will take time and be iterative until the performance of these systems is worthy of our trust. It is vital to move forward deliberately and carefully, anticipating conflicts and ethical challenges while embracing the enormous potential for improving the human condition.

## Methods

### Study Design

This prospective, single-center observational study was conducted to develop and evaluate a real-time pa-tient–data interface in a Level 4 Epilepsy Monitoring Unit (EMU) at the Hospital of the University of Pennsylvania. The study protocol was approved by the University of Pennsylvania Institutional Review Board (IRB protocol #849276), and written informed consent was obtained from all participants prior to any study procedures. Patients were enrolled during their clinical admission to the EMU, and the study period coincided with the duration of continuous video-EEG monitoring. The implementation and clinical use of artificial intelligence (AI) within this study were reviewed and approved by the Penn Medicine AI Governance Committee, which was initially established for this project and has since been institutionalized. The project aligned with the health system’s recognition of the need for a formal, expert entity to advise the IRB on patient-AI interactions in both research studies and clinical care. The committee includes representatives from informatics, information security, computer science, and clinical communities and operates under the supervision of the University IRB and Penn Medicine’s health system and School of Medicine administrations.

### Procedures

Expert epileptologists screened incoming EMU admissions to identify eligible patients. Inclusion criteria were age *≥* 18 years, intact capacity to provide informed consent and interact with an app on a smartphone, and English proficiency adequate for study procedures. Exclusion criteria were self-reported pregnancy or nursing, cognitive impairment or language barriers precluding meaningful engagement, and refusal or withdrawal of consent. Enrollment and study procedures were integrated into routine clinical care without altering standard-of-care monitoring or treatment. On the day following admission, a research coordinator approached patients to obtain informed consent and initiate enrollment. After consent was obtained, the coordinator activated the Natus (Natus, Middleton, WI) NeuroWorks software development kit (SDK) on the patient’s Natus acquisition workstation to allow real-time EEG data streaming. Each patient was provisioned an account in the study application, which included a unique username and password, data-stream linkage identifier, medical record number for medication log integration, and an EEG modality designation (scalp or intracranial) to configuration model parameters. The study application was then installed as a progressive web app on the patient’s smartphone. The patient was authenticated into their account and push notifications were enabled. The coordinator demonstrated key app features, including seizure and spike alerts, morning sleep reports, and scheduled survey questionnaires to familiarize patients with the system. Throughout their hospitalization, patients were encouraged to ask questions through the chat interface and respond to surveys and messages through the app. Patients received $25 per day of participation. The coordinator periodically checked in with the patient to discuss their experience and gather any feedback. An investigator reviewed message histories at least daily to ensure safety and appropriate behavior. Patient app accounts were deactivated on the day of discharge after completion of the exit survey. Compensation was processed after study completion, and clinical data was aggregated for research use under the approved protocol. Fig. 1 provides an overview of the study platform.

### Platform implementation

#### Overview

The platform utilized three interconnected systems (Fig. 2a). First, an on-premises data acquisition and relay layer exported real-time EEG and metadata from EMU recording stations. Second, a HIPAA-compliant cloud analytics environment ingested, preprocessed, and analyzed streaming data and persisted features and events in databases. Finally, the patient-facing app with an integrated conversational agent provided patients with clinical event notifications, surveys, and the ability to ask questions about their own physiological data. A PostgreSQL database backed the application state and messaging history, and a LangGraph-based AI agent system orchestrated data queries against a Databricks Unity Catalog via a cloud-hosted, HIPAA-secure LLM.

#### Real-time EEG acquisition and streaming

Continuous scalp or intracranial EEG signals were acquired using clinical Natus systems. Real-time access was provided by the Natus NeuroWorks SDK, which exposed ZeroMQ sockets for data, study information, and event injection on the hospital network. A cloud-hosted virtual machine subscribed to the hospital-side data publishers over ZeroMQ, retrieved sampling and channel metadata via the SDK, and batched raw samples into 1-second windows. Batches were compressed (gzip), base64-encoded, annotated with channel names, timestamps, and patient identifiers, and then forwarded to Azure Event Hubs using shared-access-signature authentication. Event Hubs fed a Databricks streaming pipeline that performed canonical EEG preprocessing, feature computation, and model inference, with outputs written to Delta tables within a Unity Catalog.

#### Cloud data processing and feature extraction

##### Preprocessing

EEG signals were bandpass filtered between 0.5-100 Hz for scalp EEG and 0.5-250 Hz for iEEG, and notch-filtered at 60 and 120 Hz to suppress powerline noise. Bad channels were identified using a validated, custom artifact rejection algorithm (Supplementary Materials). Data were then rereferenced using common average (CAR) or bipolar montages as required by specific algorithms, with bad channels excluded. Bipolar pairs rendered unavailable due to bad channels were retained but labeled as contaminated. Finally, rereferenced signals were pre-whitened. Preprocessed data at each stage, including filtering, CAR and bipolar rereferencing, prewhitening, and corresponding bad channel identifiers, were stored in a dedicated Delta Live Tables on Databricks.

##### Scalp EEG Seizure detection

For scalp recordings, we employed SPaRCNet [5], a seizure detection model capable of distinguishing among six types of ictal-interictal injury continuum (IIC) patterns and was trained on over 6,000 EEG recordings from 2,711 patients. Per model design, we applied a 10-second window with 2-second overlap on bipolar data as inputs. A segment was considered a seizure if the probability of the lateralized periodic discharges (LPD) class exceeded defined threshold. The LPD class, instead of the seizure class, was used due to higher sensitivity empirically observed, potentially due to misclassification of seizure segments as LPD. We used a data-adaptive threshold defined as the mean seizure probability plus 1.2 standard deviations, estimated from a sliding window of 400 seconds with a 100-second stride. We employed a cumulative sum strategy where a cumulative static was calculated as

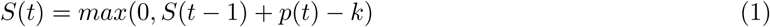

where *p*(*t*) denotes the seizure probability at time *t*, and *k* represents the adaptive threshold. A seizure detection was declared when *S*(*t*) exceeded the decision threshold (*h* = 3). The detected samples were subsequently smoothed using binary opening and closing operations. Consecutive detections separated by less than 2 seconds were merged, and merged events shorter than 10 seconds were discarded.

##### Intracranial EEG Seizure detection

For intracranial data, we evaluated both WaveNet [6] and ONCET (unpublished work) during inference and selected ONCET for fine-tuning based on its superior performance.

WaveNet was built on a WaveNet-style architecture of causal, dilating convolutional layers and trained to discriminate between seizure and non-seizure neural activity. It accepts single-channel EEG signal as input and is thus adaptable to variable channel counts in iEEG recordings. One-second segments of bipolar, prewhitened data were downsampled to 128 Hz and normalized using a robust scaler fitted on non-ictal segments prior to model input. In our implementation, the robust scaler was trained using the first 10 minutes of available data. ONCET is a seizure annotation model trained on more than a million clips of single-channel iEEG recordings of interictal, seizure onset, and seizure spread activity from 87 patients [26]. The model uses dilating residual convolution blocks with expanding receptive fields, attention, and feature compression to classify ictal activity, achieving an AUROC of 0.94 on the held-out test set and 0.95 on a gold-standard validation set [27]. ONCET inference follows similar preprocessing steps to WaveNet but operates on data downsampled to 256 Hz instead. We used a data-adaptive threshold of mean plus 0.8 standard deviations and a cumulative strategy with *h* = 3 to obtain seizure detections from each channel. A sample was classified as seizure if more than 10% of channels were predicted as seizure.

##### Spike detection

We detected scalp EEG spikes using SpikeNet [7], a convolutional neural network trained on over 13,000 annotated interictal epileptiform discharge events. SpikeNet processes 2-second windows (1-second overlap) sampled at 128 Hz, rereferenced using both bipolar (18 channels) and CAR (19 channels) montages. Predictions were generated for 1-second segments at 1-sample strides and averaged to yield a final spike probability. A threshold of 0.5 was applied for binary classification of spike presence. For iEEG, we used a separate validated spike detector [8] that processes non-overlapping 1-minute CAR data segments to output spike counts and timestamps. For both modalities, spike rate was computed as the number of detected spikes per minute. To identify periods of elevated spike activity, events separated by one minute or less were merged and segments exceeding 10 spikes per minute were defined as spike rate events.

##### Sleep staging

We used YASA [9], an automated sleep staging tool trained and validated on more than 30,000 hours of polysomnography. YASA operates on a single central EEG channel (C3, C4, or Cz) referenced to M1 or Fpz, downsampled to 100 Hz, and band-pass filtered (0.4–30 Hz). EEG segments of at least five minutes are classified into wake, REM, N1, N2, or N3 sleep stages in 30-second intervals. In our implementation, we applied YASA to non-overlapping 10-minute windows to improve prediction stability. Sleep stages were then smoothed with a mode filter (window size 15). When central channels were unavailable, we computed alpha-delta ratio as an alternative measure. Using non-overlapping 1-minute windows applied to the CAR data, we calculated the ratio of mean alpha power (8–13 Hz) to mean delta power (1–4 Hz) averaged across channels. Sleep events were defined by grouping consecutive samples of non-wake sleep stages, allowing gaps of up to 1.5 minutes and discarding events shorter than five minutes. Auto-generated sleep reports summarized the absolute duration and relative proportion of each sleep stage in 12-hour periods (19:00–07:00 and 07:00–19:00).

##### Synchrony

To quantify brain-wide phase synchronization, we used non-overlapping 1-minute windows from the CAR data. The analytic signal was derived via the Hilbert transform to extract instantaneous phases. The degree of phase synchrony across non-artifact channels was computed at each time point and averaged over time to create a global synchrony index.

##### Anti-seizure medication load

We estimated total anti-seizure medication plasma concentrations from medication dosing records using a previously validated first-order pharmacokinetic model published recently by our group [28]. Medication records were retrieved from Sickbay (Medical Informatics Corp., Houston, TX, USA) and updated every five minutes, with anti-seizure medication load recalculated in 1-minute intervals.

##### Visualization

Identified events were visualized to facilitate interpretation and potential interaction with patients. For scalp seizure events, the absolute slope of the EEG signal was calculated for each channel as a measure of activity. These data were visualized as a heatmap over time and as animated head topographic maps. For iEEG seizure events, predicted seizure probabilities across channels were additionally plotted as heatmaps. Sleep stages were visualized as hypnograms over 12-hour periods, spanning 19:00 to 07:00 and 07:00 to 19:00. Spike rates were visualized as line plots. Fig. 1 shows example figures from each modality as they were presented to patients, depending upon how they queried the AI agent.

### Patient-facing application

The application was a containerized Python Flask service served by Gunicorn and deployed to Azure App Service. It provided authentication, a chat interface, and push notifications (web push protocol with VAPID). A dedicated Azure Database for PostgreSQL stored patient authentication information, conversations, surveys, synchronized seizure and spike events, and cached images linked from Databricks volumes. A background scheduler (APScheduler) polled Databricks tables at 30s intervals to synchronize new events and dispatch daily sleep reports once morning surveys were completed. To restrict patient interactions to high-confidence detections, the system was configured to poll the database only for events meeting certain thresholds. Specifically, only scalp events with mean probability *≥* 0.80 and maximum probability *≥* 0.95, and iEEG events exceeding one minute in duration were retrieved. Push notifications for clinical events were disabled overnight (21:00–08:00), but these events remained accessible to patients via data queries. Images (e.g., sleep staging hypnograms and seizure heatmaps) were retrieved from Databricks volumes via the Files API, cached under application-controlled storage, and displayed in-line within the chat history.

The patient application and cloud analytics environment were linked via three bidirectional data flows: (1) patient-initiated queries with secure SQL filtering delivered personalized clinical event summaries (e.g., historic seizure characterization) back to the chat, (2) system-initiated alerts synchronized detection events from the analytics environment to the application, triggering push notifications, and (3) patient-in-the-loop annotations captured patient responses to these events, which were persisted back in Databricks for iterative model retraining.

### Conversational agent

The conversational agent (Fig. 2b) was implemented as a directed state machine using LangGraph, with nodes for message moderation, routing based on classified patient intent (general chat vs. data agent query), conditional message evaluation and clarification, and a ReAct-style data agent with an iterative tool loop [29]. The persistent state maintained conversation history, patient context (unique identifier, EEG type, sleep-monitoring status), routing flags, moderation results, and SQL query metadata. All SQL queries were executed through a custom tool that injected a patient-specific filter for data isolation and to match database table semantics. To manage the context length of tokens input to the LLM, images were pruned from chat history when not required and replaced with descriptive placeholders. The agent had a bounded iteration loop (up to 25 tool calls) and returned an automated message on recursion or validation failures.

### Surveys

The application administered three daily surveys: morning (08:00–09:00), afternoon (14:00–15:00), and evening (20:00–21:00), as well as a one-time exit survey at study completion. Delivery times were randomized per patient within each window to reduce anticipatory bias while maintaining general daily timing. Each survey contained 10 questions (0–100 visual analog scale with labeled endpoints); two for each of the following five constructs: depressed mood, anxiety, perceived cognitive function, stress level, and fatigue/energy level. The morning survey included an additional five items from the Richards Campbell Sleep Questionnaire [30] to assess sleep depth, latency, wakefulness, maintenance, and overall quality. The exit survey comprised the 10-item System Usability Scale [12] (SUS; 5-point Likert) and four free-text questions on system utility and patient feedback. After survey completion, follow-up questions were generated under pre-specified conditions: a fixed, open-ended follow-up after morning surveys regarding factors affecting sleep, and up to three personalized follow-ups after afternoon/evening surveys when responses exhibited significant day-to-day changes, monotonic trends, or sustained extremes, with at most one follow-up per construct. Follow-up question content was generated by an LLM and they expired after one hour. Surveys themselves expired if not completed within three hours after being administered. Full survey questionnaires are provided in the Supplementary Materials.

### Interaction and engagement analysis

To quantify patient engagement and system performance, we calculated response latencies for both the AI agent system and patients. System response latency was defined as the time difference between a patient’s message and the subsequent AI response. For survey follow-up questions generated by the LLM, latency was calculated from the timestamp of survey completion to the delivery of the first follow-up question. Patient response latency was categorized by interaction type. For general chat, latency was measured from the delivery of an AI message to the first subsequent patient message. For surveys, latency was defined as the time from notification delivery to the patient initiating the survey. Similarly, for seizure and spike events, latency was calculated from the time of notification delivery to the patient’s annotation of the event. Prior to analysis of the chat interface data, we applied exclusion criteria to the message logs that removed automated (non-AI-generated) system messages.

## Model fine-tuning

### Patient-in-the-loop fine-tuning

We implemented real-time model fine-tuning every three hours separately for each patient. Patient annotations were converted into binary labels time-aligned with model predictions, where a window was labeled as 1 if it overlapped with any patient-confirmed seizure event and 0 otherwise. Model-detected events without corresponding annotations were labeled as NaN and excluded from fine-tuning. Fine-tuning was skipped for any 3-hour session with no detected events or when all detected events lacked valid annotations.

Samples were split into training and validation sets with an 85:15 ratio. To preserve representation of rare positive cases, we adopted a stratified temporal split in which the first 85% of seizure and non-seizure samples were assigned to the training set and the remaining 15% to the validation set. Fine-tuning was also skipped if the validation AUROC exceeded 0.98, indicating near-perfect performance, or was below 0.5, suggesting potentially incorrect patient annotations. For SPaRCNet, the Kullback–Leibler (KL) divergence over the six-class output distribution was used as loss. For ONCET, channel-level seizure probabilities were aggregated using a top-*k* pooling strategy, as only sample-level labels (without channel-wise annotations) were available. Given channel probabilities 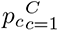, the aggregated probability, defined as

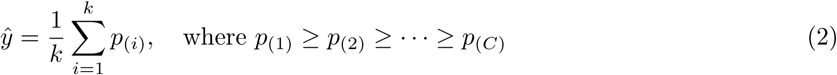

denotes the sorted channel probabilities in descending order, and *k* was set to 10% of the total number of channels. A binary cross-entropy loss was then computed between the ground-truth label and ŷ. Given the substantial class imbalance toward non-seizure samples, we incorporated model distillation to mitigate collapse to the negative class by introducing an additional supervision loss defined as the KL divergence between the original model’s predictions and those of the fine-tuned model, weighted by a factor of 0.5.

We used an optimizer with a learning rate of 5 *×* 10*^−^*^6^ and weight decay of 1 *×* 10*^−^*^3^. A weighted random sampler was employed to oversample false-positive, false-negative, and true-positive samples according to the inverse frequency of each class. For SPaRCNet, we additionally applied channel-flipping augmentation to reduce overfitting by duplicating each EEG segment with left and right channels swapped. During fine-tuning, only a subset of layers was set as trainable, while the remaining layers were frozen to avoid catastrophic forgetting. For SPaRCNet, the initial convolutional layer, the final dense block, and the fully-connected layers were trainable. For ONCET, the feature compression and final classification layers were set as trainable. We further implemented a progressive layer-unfreezing schedule: if performance did not improve for 2 consecutive epochs, an additional block or layer was unfrozen.

Each fine-tuning session was trained for up to 20 epochs. In addition to training and validation loss, AUROC, AUPRC, and other performance metrics were monitored. Model checkpoints were saved whenever the validation loss improved or when both AUROC and AUPRC improved relative to the previous best model. Early stopping was applied with a patience of 3 epochs for SPaRCNet and 10 epochs for ONCET, and training was also terminated if either AUROC or AUPRC exceeded 0.98.

### Clinician-in-the-loop fine-tuning

Deep active learning was used to identify the most informative portions of each day’s EEG recording and present them to clinicians for annotation, enabling continuous refinement of the detection model with efficient expert input. At the end of each day, the model performed inference on the full day’s recording. Using a least-confidence sampling strategy, the system selected ten 90-second segments for which the model showed the greatest uncertainty. These segments were sent to clinicians for review and labeling. The clinician-annotated segments were then used to fine-tune the model. During fine-tuning, the backbone was partially unfrozen using the same configuration in the patient fine-tune, and model distillation was applied with a weighting factor of 0.5. After fine-tuning, the updated model was deployed for seizure detection on the subsequent days’ recordings.

### Performance Evaluation

We evaluated the performance of fine-tuned models post hoc to determine whether fine-tuning improved seizure detection on subsequent data. We first performed a validation using patient-initiated push-button alarms in the EMU, instead of patient responses through the system. Alarms were converted to annotations for fine-tuning, with the period from one minute before to two minutes after each alarm as a seizure event, and all remaining periods as non-ictal. This approach enabled evaluation in a larger cohort, including patients recruited before the interactive seizure-annotation feature was available. Seizures annotated by EMU clinicians were used for clinician-in-the-loop fine-tuning and served as the ground truth. All annotations were converted into binary labels time-aligned with model predictions. The model was fine-tuned sequentially at multiple time points, and its performance on subsequent data not used for fine-tuning was compared to the baseline (Fig. 5a). We also evaluated performance across the entire EMU stay by applying the fine-tuned models once they became available and comparing to the baseline. Multiple performance metrics were computed using the SzCORE framework [31], including event-level seizure sensitivity, false alarms per hour, and event-wise F1 score. During evaluation, seizure events separated by less than 90 seconds were merged, and events longer than 5 minutes were split into separate events. A seizure event was considered detected if any predicted event overlapped with it, with a 30-second preictal tolerance and a 60-second postictal tolerance. AUROC and AUPRC were also calculated.

We next evaluated prospective fine-tuning using patient-provided annotations collected through the system. For safety reasons, only the validated baseline model was deployed during actual patient sessions to avoid issuing inaccurate alarms that could affect the patient. To approximate real-time application, we performed a pseudo-prospective analysis in which the model was sequentially fine-tuned, mirroring the process of live deployment. Patient annotations were converted to labels as described above, and all other evaluation procedures followed the same protocol as in the validation analyses.

### Statistical analysis

For each fine-tuned model, we computed the performance difference relative to the baseline model on future test data (Fig.5a). The median performance difference across all fine-tuned models was calculated for each patient. These medians were compared to zero using a one-sided Wilcoxon signed-rank test, with the alternative hypothesis set according to the expected direction of effect. Performance across the EMU stay was compared between the baseline and fine-tuned models using a one-sided paired Wilcoxon signed-rank test. To assess the association between synchrony and anti-seizure medication load, we applied a linear mixed-effects model using restricted maximum likelihood estimation. Total anti-seizure medication load was treated as the dependent variable, with synchrony and alpha–delta ratio included as fixed-effect predictors and patient as a random effect. To compare system response latencies between the general chat and data agent interaction modes, we used a two-sided Mann-Whitney U test. A rank-biserial correlation was computed as a measure of effect size.

## Data Availability

All data produced in the present study are available upon reasonable request to the authors.

## Acknowledgments

This work was funded by an NIH Pioneer Award, DP1-NS-122038, NIH/NINDS R01-NS-125137, NIH T32NS091006, foundation support from Jonathan and Bonnie Rothberg, Neil and Barbara Smit, and the Small Lake Foundation. K.A.D. received funding from NIH/NINDS, awards no. R61-NS-125568 and R01-NS-116504. N.S. received funding from NIH/NINDS, award no. K99NS138680, and the Department of Defense, grant no. W81XWH2210593. E.C. received support from NIH/NINDS, award no. K23 NS121401-01A1 and the Burroughs Wellcome Fund. We are grateful for all patients and their families for their involvement in the research program. We thank L. Faillace, L. Simmons, and the staff, fellows, and technicians of the Hospital of the University of Pennsylvania epilepsy monitoring unit for their invaluable assistance and cooperation during the study. We also thank S. McCurdy and J. Data for their help in setting up the real-time EEG data streaming service and D. Lydon-Staley for providing survey expertise.

## Supplemental Materials

### Supplementary methods

#### EEG artifact rejection

For each one-second EEG segment, we detected bad channels using a custom algorithm. Channels were considered bad and excluded from further analyses if: (1) more than 50% of samples were missing or zero-valued, (2) more than 50% of samples were flat-lined, (3) absolute deviations from the channel-wise median exceeded 5 mV for more than 10 samples, (4) extreme outliers were present, defined as values falling outside the envelope defined by the median *±*10 *×* (99th percentile - median), consistent with transient disconnection or motion artifacts, and (5) power in the 58–62 Hz band exceeded 10% of total spectral power.

## Supplementary figures

**Figure S1:**
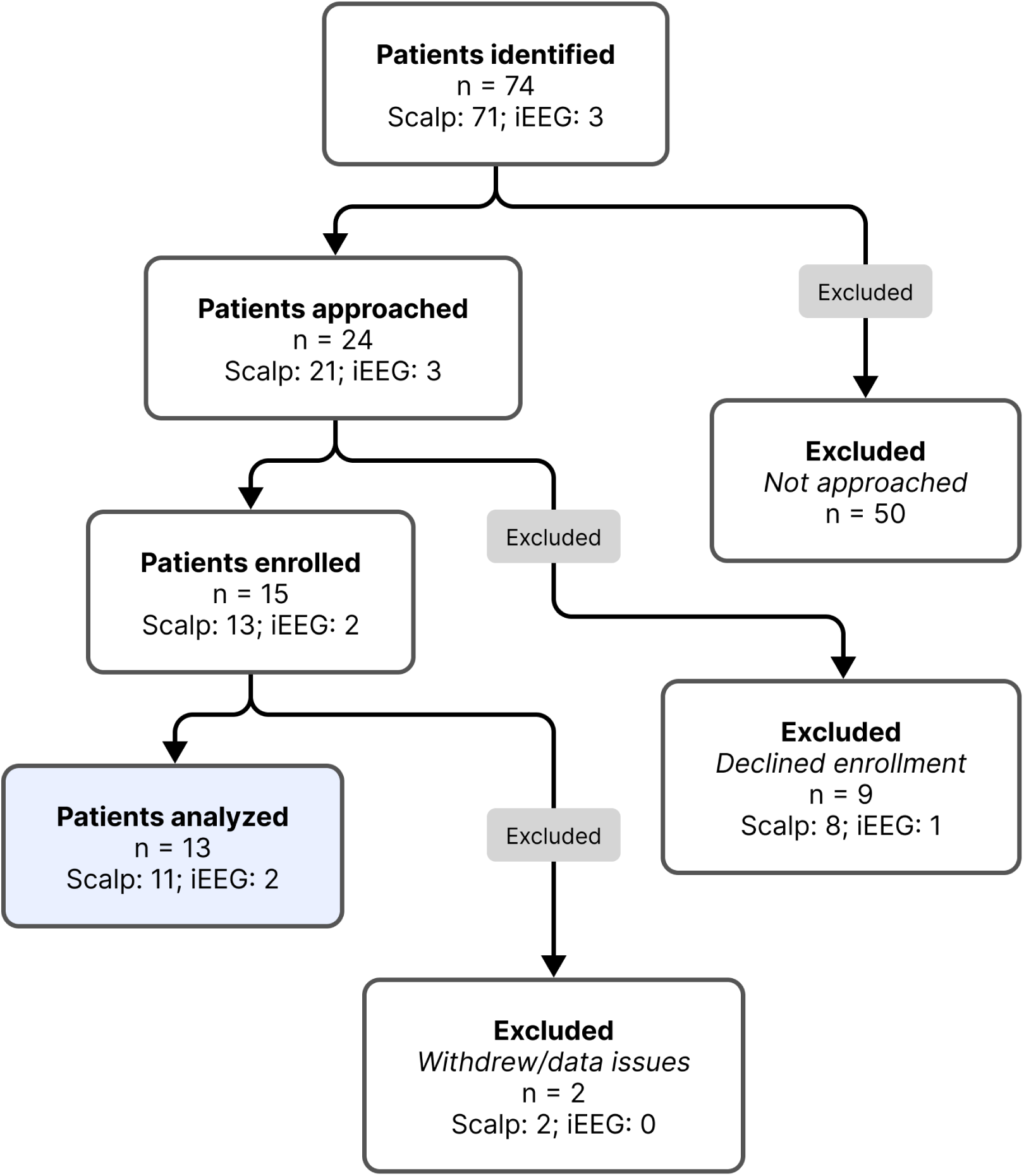
Patient enrollment and analysis flowchart.

**Figure S2:**
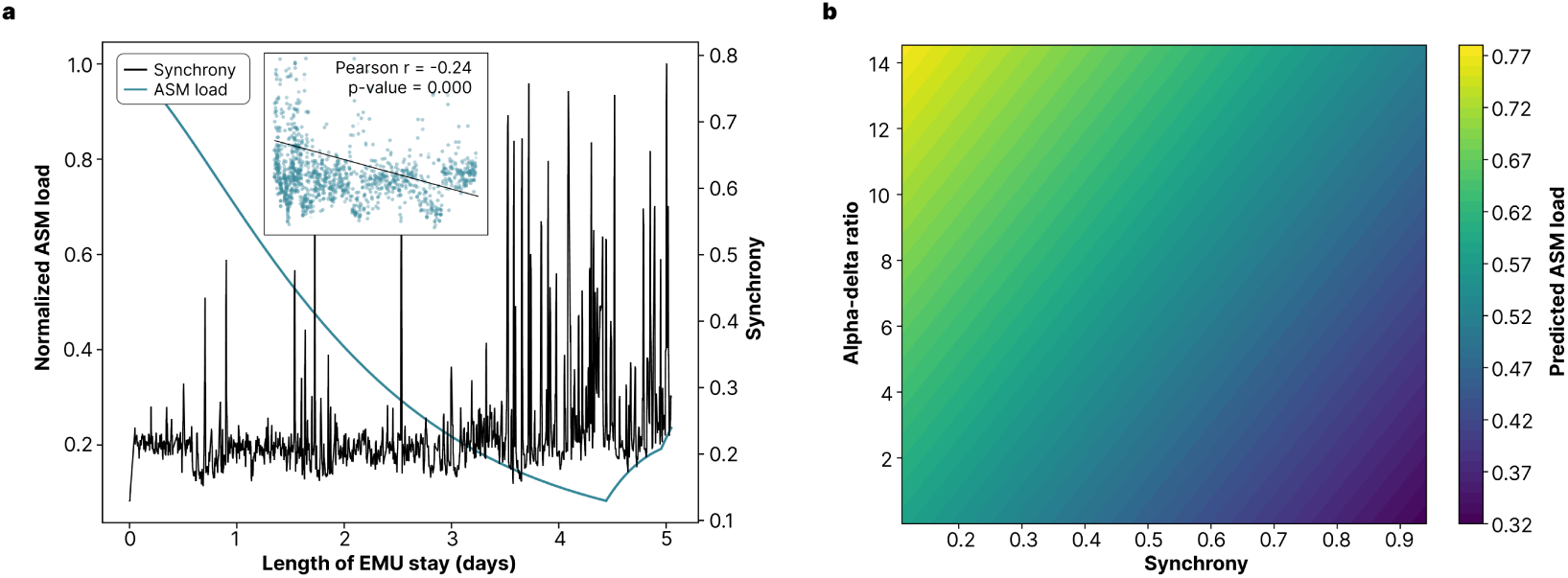
Synchrony as a biomarker for anti-seizure medication load. **a.** Time series of synchrony and normalized anti-seizure medication (ASM) load across the entire epilepsy monitoring unit (EMU) stay for a representative patient. **b.** Predicted response surface from a linear mixed model with normalized ASM load as the dependent variable, synchrony and alpha–delta ratio as fixed-effect predictors, and patient as a random effect.

**Figure S3:**
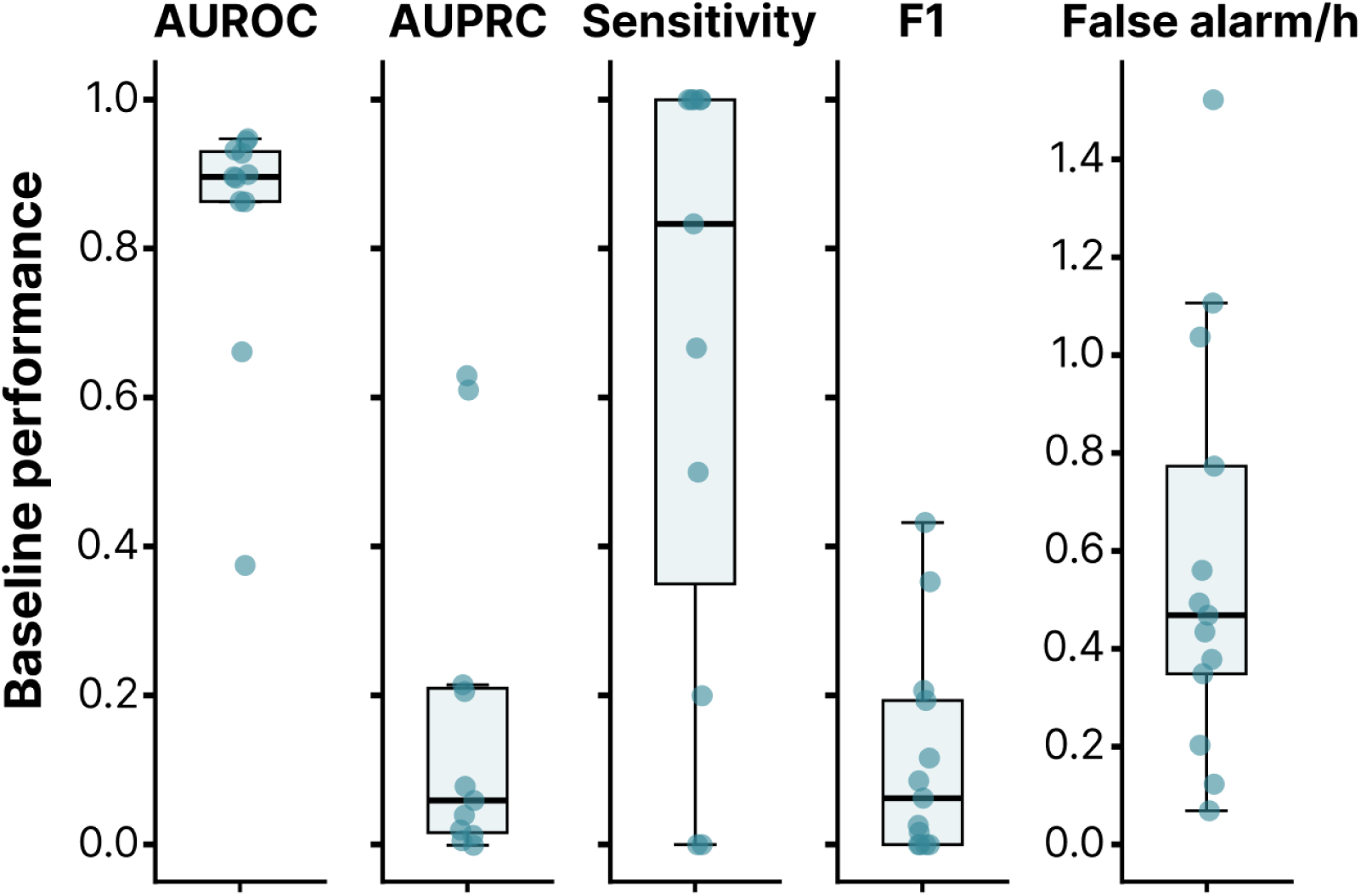
Seizure detection performance at baseline. Each dot represents one patient. The box represents the interquartile range (IQR) with a central line indicating the median across patients; whiskers extend to 1.5×IQR. **Abbreviations:** AUROC: area under the receiver operating characteristic curve; AUPRC: area under the precision-recall curve.

## Supplementary tables

**Table S1:** Summary of quantitative EEG features and detection algorithms.

| Domain | Feature/model | Window | Modality<sup>a</sup> | Preprocessing requirement | Missing data handling |
| --- | --- | --- | --- | --- | --- |
| Sleep | Alpha-delta ratio | 1 min | Both | CAR | Discard invalid |
| Sleep | YASA | 10 min<sup>b</sup> | Both<sup>c</sup> | CAR | Discard if <5min valid |
| ASM estimation | Synchrony | 1 min | Both | CAR | Drop window |
|  | ASM load | 1 min | Both | N/A | N/A |
| Spike detection | SpikeNet | 2s (1s shift) | Scalp | CAR and bipolar | Drop window |
|  | Spike detector | 1 min | iEEG | CAR | Drop window |
| Seizure detection | SPaRCNet | 10s (2s shift) | Scalp | Bipolar | Drop window |
|  | WaveNet | 1s | iEEG | Bipolar prewhitened | Drop window |
|  | ONCET | 1s | iEEG | Bipolar prewhitened | Drop window |

**Abbreviations:** CAR: common average re-reference; Bipolar: bipolar re-reference; YASA: Yet Another Spindle Algorithm; ASM: anti-seizure medication; iEEG: intracranial EEG.
<sup>a</sup> Whether the feature or model is applied on scalp or intracranial EEG.
<sup>b</sup> Calculated at 30s intervals.
<sup>c</sup> Requires scalp channels C3, C4, or Cz.

**Table S2:** Event definitions and post-processing criteria.

| Event | Source model | Criteria; Additional metrics |
| --- | --- | --- |
| Seizure detection | SPaRCNet (scalp) | Duration $\geq 10s$ (gap $\leq 2s$); Mean prob $\geq 0.8$, max $\geq 0.95$ |
| | WaveNet (iEEG) | Duration $\geq 10s$ (gap $\leq 2s$) |
| | ONCET (iEEG) | Duration $\geq 10s$ (gap $\leq 2s$) |
| Elevated spike rate | SpikeNet (scalp)<br>Spike detector (iEEG) | Duration $\geq 1$ min (gap $\leq 1$ min); Mean rate $\geq 10$ spikes/min |
| Sleep report | YASA | 12h blocks (7:00–19:00 / 19:00–7:00); Absolute duration and proportion of each stage |

**Abbreviations:** iEEG: intracranial EEG; YASA: Yet Another Spindle Algorithm.

**Table S3:**
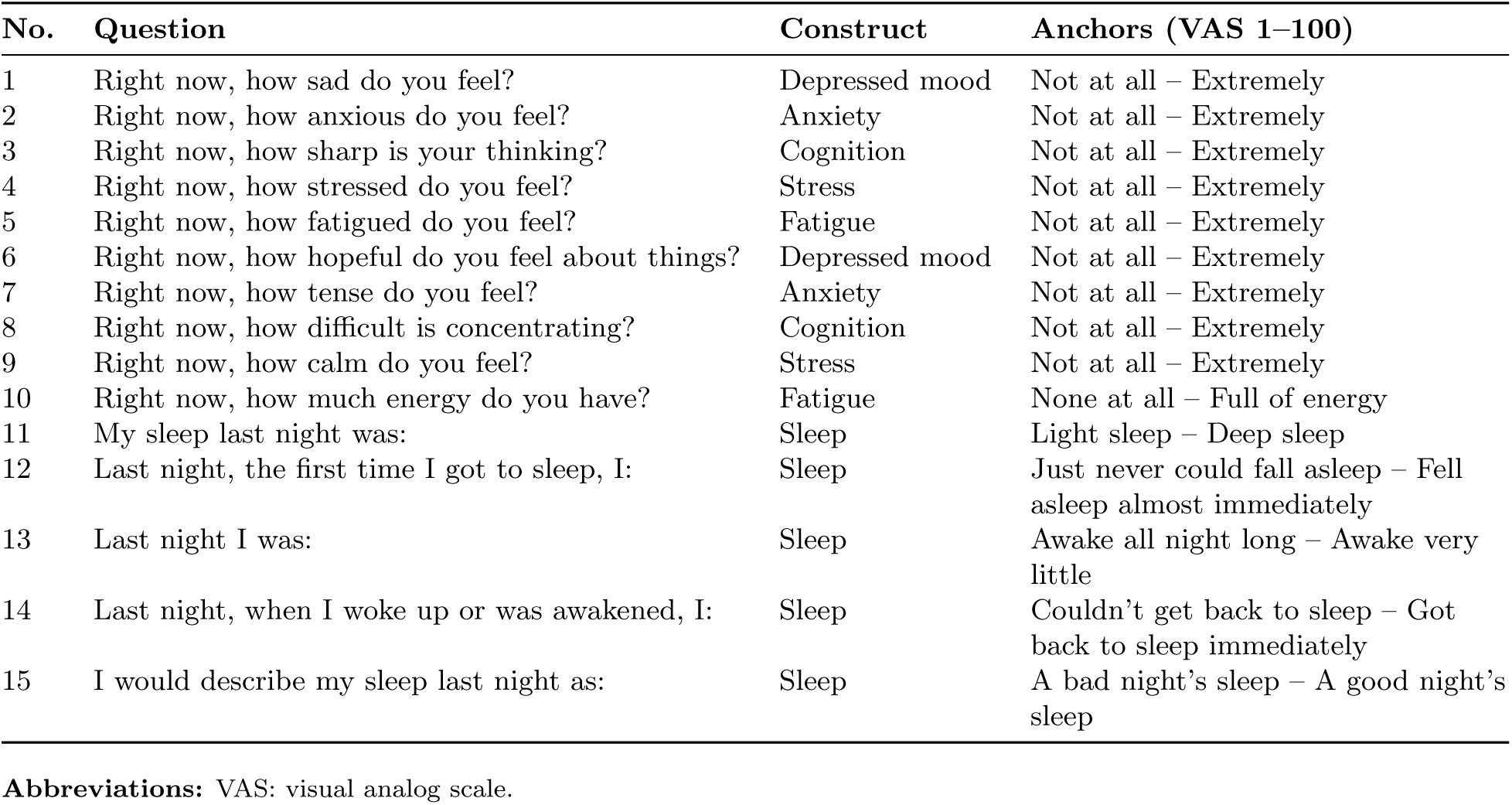
Morning survey (8:00–9:00).

**Table S4:**
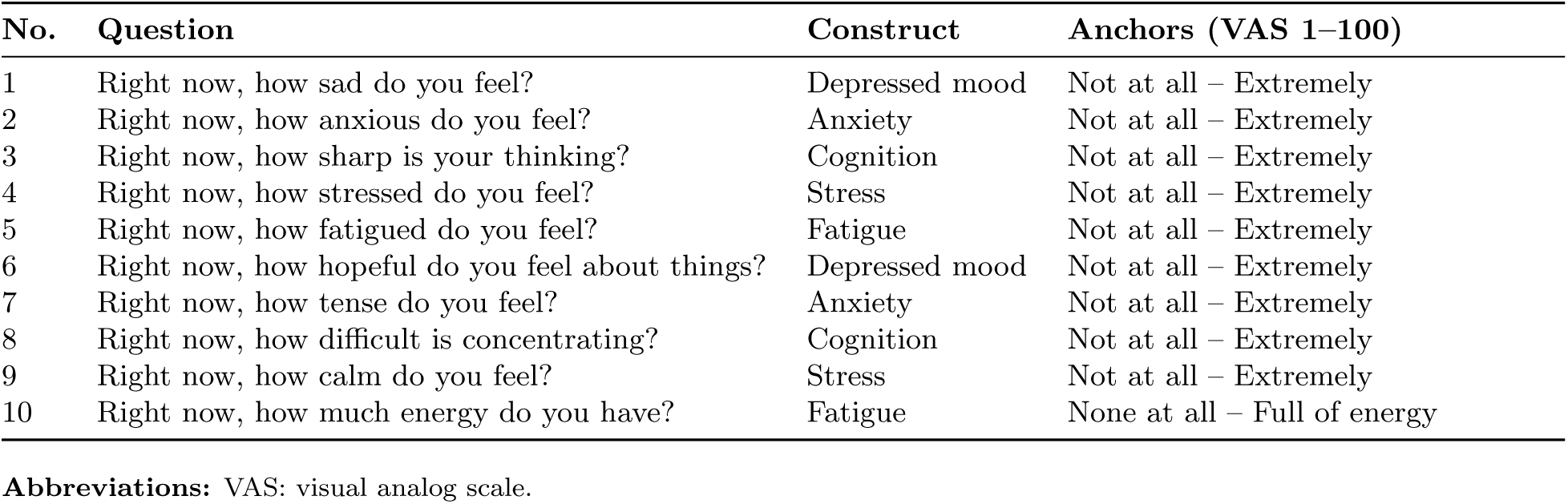
Afternoon survey (14:00–15:00).

**Table S5:**
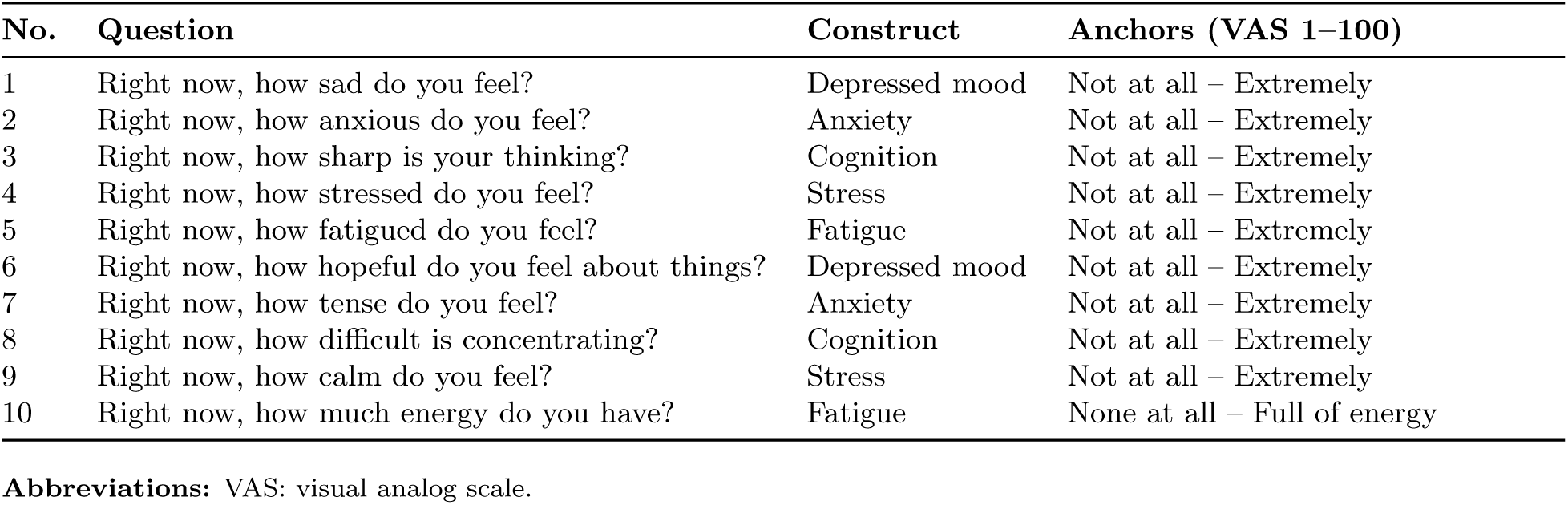
Evening survey (20:00–21:00).

**Table S6:**
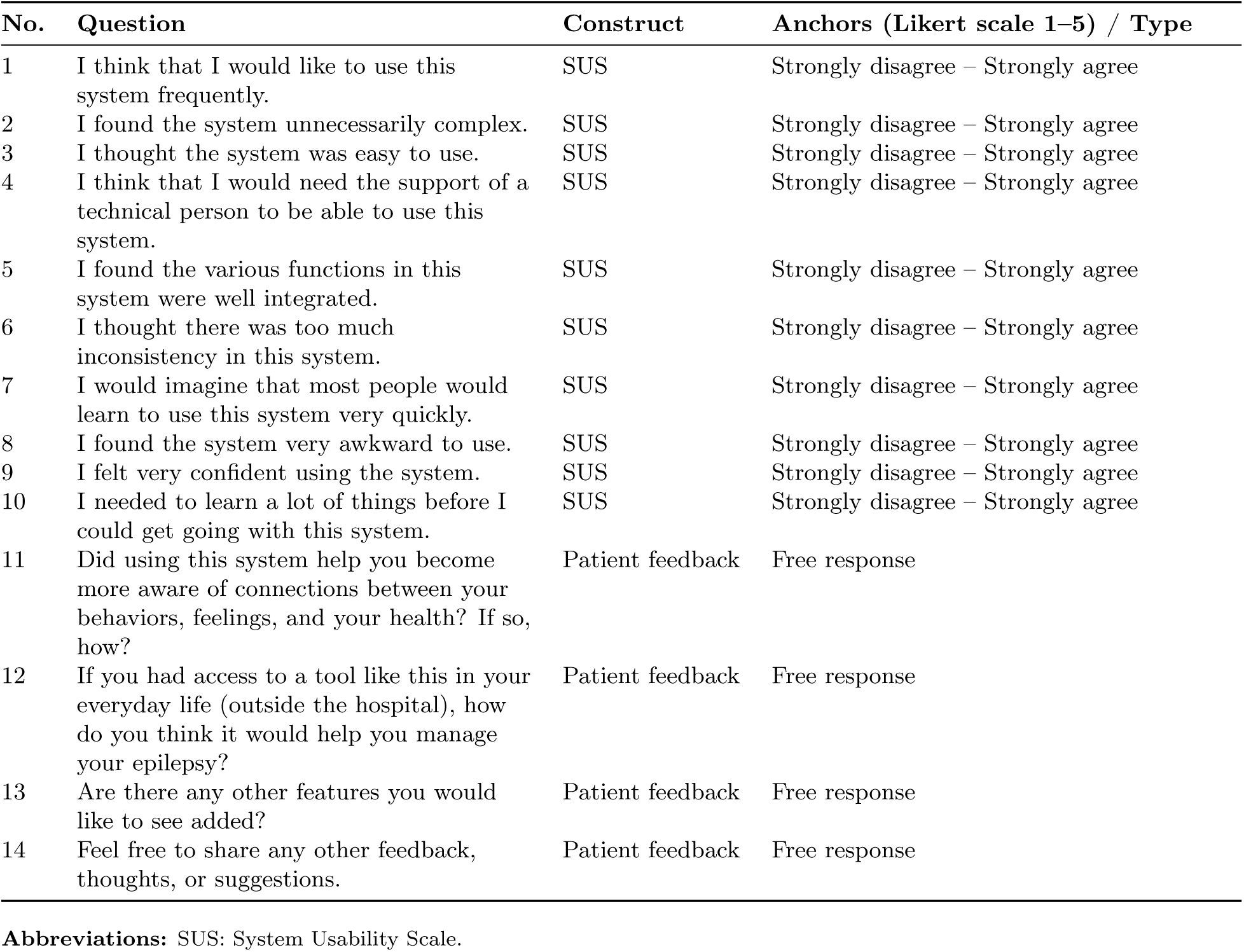
Exit survey.

## References

[1] Begley, C. et al. The global cost of epilepsy: A systematic review and extrapolation. Epilepsia 63, 892–903 (2022).

[2] Jette, N. & Wiebe, S. Update on the surgical treatment of epilepsy:. Current Opinion in Neurology 26, 201–207 (2013).

[3] Nair, D. R. et al. Nine-year prospective efficacy and safety of brain-responsive neurostimulation for focal epilepsy. Neurology 95 (2020).

[4] Rehman, M., Higdon, L. M. & Sperling, M. R. Long-Term Home EEG Recording: Wearable and Implantable Devices. Journal of Clinical Neurophysiology 41, 200–206 (2024).

[5] Jing, J. et al. Development of Expert-Level Classification of Seizures and Rhythmic and Periodic Patterns During EEG Interpretation. Neurology 100, e1750–e1762 (2023).

6. Revell, A. Y. et al. A Taxonomy of Seizure Spread Patterns, Speed of Spread, and Associations With Structural Connectivity. Tech. Rep., Neuroscience. URL http://biorxiv.org/lookup/doi/10.1101/2022.10.24.513577.

[7] Jing, J. et al. Development of Expert-Level Automated Detection of Epileptiform Discharges During Electroencephalogram Interpretation. JAMA neurology 77, 103–108 (2020).

[8] Aguila, C. A. et al. Mesial-to-lateral gradients of epileptiform activity to localize mesial temporal lobe epilepsy. Epilepsia 66, 3491–3504 (2025).

[9] Vallat, R. & Walker, M. P. An open-source, high-performance tool for automated sleep staging. eLife 10, e70092 (2021).

10. Ma, D., et al. Synchrony is a robust iEEG biomarker for antiseizure medication load in epileptic patients (2026).

[11] Ung, H. et al. Interictal epileptiform activity outside the seizure onset zone impacts cognition. Brain 140, 2157–2168 (2017).

12. Brooke, j. SUS: A ’Quick and Dirty’ Usability Scale. In Usability Evaluation In Industry (CRC Press, 1996).

[13] Bernini, A., Dan, J. & Ryvlin, P. Ambulatory seizure detection. Current Opinion in Neurology 37, 99–104 (2024).

[14] Beran, R. G. Use of Interval Therapy with Benzodiazepines to Prevent Seizure Recurrence in Stressful Situations. Brain Sciences 12, 512 (2022).

[15] Snyder, D. E., Echauz, J., Grimes, D. B. & Litt, B. The statistics of a practical seizure warning system. Journal of Neural Engineering 5, 392–401 (2008).

[16] Cook, M. J. et al. Prediction of seizure likelihood with a long-term, implanted seizure advisory system in patients with drug-resistant epilepsy: A first-in-man study. The Lancet Neurology 12, 563–571 (2013).

[17] Saab, K., Dunnmon, J., Ré, C., Rubin, D. & Lee-Messer, C. Weak supervision as an efficient approach for automated seizure detection in electroencephalography. npj Digital Medicine 3, 59 (2020).

[18] Roth, D. Incidental Supervision: Moving beyond Supervised Learning. Proceedings of the AAAI Conference on Artificial Intelligence 31 (2017).

[19] Xu, Z. et al. Annotating neurophysiologic data at scale with optimized human input. Journal of Neural Engineering 22, 046003 (2025).

[20] Pearson, C. et al. Care access and utilization among medicare beneficiaries living with Parkinson’s disease. npj Parkinson’s Disease 9, 108 (2023).

[21] Team, G. et al. Gemma 3 technical report (2025). URL https://arxiv.org/abs/2503.19786.2503.19786.

22. Abdin, M., et al. Phi-3 Technical Report: A Highly Capable Language Model Locally on Your Phone (2024). 2404.14219.

23. Wu, K., Zhao, Z. & Yener, B. Large EEG-U-Transformer for Time-Step Level Detection Without Pre-Training (2025). 2504.00336.

[24] Sverrisson, Þ. & Guðmundsson, S. LookAroundNet: Extending Temporal Context with Transformers for Clinically Viable EEG Seizure Detection (2026). 2601.06016.

[25] Yao, Y., Wang, H., Chen, L., Peng, Y. & Luo, J. Foundation models for EEG decoding: Current progress and prospective research. Journal of Neural Engineering 22, 061002 (2025).

[26] Ojemann, W. K. S. et al. Unsupervised seizure annotation and detection with neural dynamic divergence. medRxiv.

[27] Ojemann, W. K. S., et al. Multi-expert consensus annotations of spontaneous and stimulation- induced seizures in stereotactic EEG. medRxiv (2026).

[28] Ghosn, N. J. et al. A pharmacokinetic model of antiseizure medication load to guide care in the epilepsy monitoring unit. Epilepsia 64, 1236–1247 (2023).

29. Yao, S., et al. ReAct: Synergizing Reasoning and Acting in Language Models (2023). Comment: v3 is the ICLR camera ready version with some typos fixed. Project site with code: https://react-lm.github.io, 2210.03629.

[30] Richards, K. C., O’Sullivan, P. S. & Phillips, R. L. Measurement of Sleep in Critically Ill Patients. Journal of Nursing Measurement 8, 131–144 (2000).

[31] Dan, J. et al. SzCORE : Seizure Community Open-Source Research Evaluation framework for the validation of electroencephalography -based automated seizure detection algorithms. Epilepsia 66, 14–24 (2025).

